# Turning Lung Clearance Index on its head. Reference data for SF_6_ Multiple Breath Washout Derived Ventilation distribution efficiency (VDE)

**DOI:** 10.1101/2022.09.11.22279825

**Authors:** Rikke Mulvad Sandvik, Anders Lindblad, Paul D Robinson, Kim G Nielsen, Per Gustafsson

## Abstract

**Introduction:** Cystic fibrosis (CF) is characterized by increased ventilation inhomogeneity (VI), as measured by multiple breath washout (MBW), from infancy. Lung clearance index (LCI) is the most reported VI outcome. This study aimed to evaluate historically published reference equations for sulphur hexafluoride (SF_*6*_) MBW outcomes, to data collected using updated commercial SF_*6*_MBW equipment and to produce device specific equations if necessary.

**Method:** SF_*6*_MBW was performed in 327 healthy children aged 0.1-18.4 years (151 [46%] girls), 191 (58.4%) < 3 years. Z-scores were calculated from published reference equations (FRC and LCI) and multivariate linear regression performed to produce device specific reference equations. Due to increasing residual standard deviations with increasing LCI values, investigation of methods for improvement, were investigated, based on the relationship between VI and dead space ventilation (VD/VT; dead space volume/tidal volume) in a cohort of 59 healthy children, 26 children with CF (n=138 test occasions) and 49 adults with lung disease.

**Results:** Historical SF_*6*_MBW reference equations were unsuitable for Exhalyzer D® data. In contrast to LCI and log_10_(LCI), 1/LCI (ventilation distribution efficiency; VDE) was linearly related to VD/VT, with z-scores linearly related to its absolute values. Reference equations were reported for VDE and log_10_(FRC). Significant predictors for VDE and log_10_(FRC), respectively, were log_10_(age) and gender, and log_10_(height), gender and posture.

**Conclusion:** 1/LCI (e.i., FRC/CEV[%]) reflecting ventilation distribution efficiency (VDE) in the lungs was a superior index of ventilation inhomogeneity compared to LCI and log_10_(LCI) due to its linear relationship to VD/VT.

## BACKGROUND

Multiple-breath washout (MBW) is a well-established method (1) for determination of resting lung volume (functional residual capacity, FRC) and the clearance rate of an inert marker gas to reflect ventilation inhomogeneity (VI). The latter is most commonly reported as the lung clearance index (LCI) – first described by Margaret Becklake in 1951(2). LCI, as used today, is calculated as the ratio between the cumulative expired volume (CEV) of air required for reducing end-tidal concentration of an inert marker gas to 1/40 ^th^ of its starting concentration, and the simultaneously measured FRC: LCI = CEV/FRC. Importantly, LCI has been demonstrated to be a sensitive marker of lung involvement in cystic fibrosis (CF) (3). Requiring only tidal breathing and passive cooperation, the MBW method is feasible across the entire paediatric age range, even in infancy (4–6), where the emergence of detectable impairment has been shown to occur (7,8). This utility in young children highlights the importance of robust age-appropriate reference data starting from infancy. Defining pediatric MBW reference equations including the first two years of life is challenging because VI changes over time due to respiratory system development (9–12), including rapid alveolarization, which occurs with important gender differences leading to a greater number of alveoli in boys and a larger alveolar surface area than girls for a given age and stature(9). Improvement in LCI (i.e. decreasing values) has been confirmed in two sulfur hexafluoride (SF_*6*_) MBW studies of healthy young children (aged 2 weeks to 19 years, and 3-60 months, respectively) using the gold standard respiratory mass spectrometer (RMS) based MBW system (13,14). Since 2016 the availability of a commercial SF_*6*_MBW system suitable for use in all age groups (Exhalyzer D®) has emerged (4–6,15,16), but applicability of the previously published RMS SF_*6*_MBW derived reference equations to this equipment(13,14) remains unclear (see Appendix for details of device differences).

### Study aims

The primary aims of this study were to i) evaluate the relevance of historical reference equations for mass spectrometer derived SF_*6*_MBW outcomes (FRC and LCI) (13,14) to data collected using the commercial EXHALYZER D® SF_*6*_MBW system in a contemporaneous large cohort of healthy infants, toddlers and older children, and ii) produce new device specific reference equations if necessary. Due to the challenges, we faced in producing these new reference equations and the relationships exposed between LCI and dead space ventilation during our evaluation, we explored alternate approaches to expressing VI based on LCI to avoid the issues revealed. We assessed the physiological applicability of our improved index of ventilation distribution efficiency (VDE) in an additional cohort containing both health and disease to generate a greater range of VI. That process is described in the text that follows.

## METHOD

### Design

This was a cross-sectional, international, collaborative study between three centers: Department of Pediatrics, General Hospital, Skövde, Sweden; Cystic Fibrosis Centre; Queen Silvia Children’s Hospital, Gothenburg, Sweden; and CF Centre Copenhagen, Danish Paediatric Pulmonary Service, Department of Paediatric and adolescent medicine, Copenhagen University Hospital.

### Ethics

Ethics Committee approval was obtained from the University of Gothenburg, Sweden (DNR 746-15) for the Swedish data collection and from the Ethics Committee of Capital Region of Denmark (H-18057001) for the Danish data collection. All parents provided written informed consent prior to participation in the study.

### Study participants for SF_6_MBW reference equations derivation

Two groups of healthy children were recruited for this cross-sectional study. Group 1 (tested supine during sleep) targeted healthy term-born (gestational age ≥37.0 weeks) infant/toddlers (age 0-36 months, < 15kg) from both Sweden and Denmark, and group 2 (tested awake upright sitting) targeted healthy term-born preschool and school aged children (age ≥3 years) from Sweden, only. All children were tested at a time free of any respiratory tract symptoms (for ≥1 week in infants/toddlers and ≥3 weeks in older subjects). Exclusion criteria were history of any lung diseases (including asthma or bronchiolitis), or malformations or diseases to respiratory system, heart, liver, kidney, or nervous system, from birth to study visit.

### MBW recordings and quality control

Triplet MBW recordings, using a mixture of 4.0% SF_*6*_ in air for wash-in until equilibration, were performed with the EXHALYZER D® (ECO MEDICS AG, Duernten, Switzerland) using SPIROWARE® 3.2.1. Off-line breath by breath quality control analysis, as described in several previous publications (4,6,15,16), was performed using an in-house software developed in LabVIEW® (National Instruments, Austin, TX) based on identical algorithms used in the SPIROWARE® software. Group 1 performed MBW in the supine position, during quiet sleep (natural or after light sedation with intranasal Dexmedethomidine), with the EXHALYZER D® infant set-up, dead space reducer #1, and a putty sealed Rüsch face mask #1 or #2. Mask dead space was estimated at 2-7 mL and not corrected for in the analysis. Group 2 performed MBW awake, upright sitting, with the dead space reducer #2 or #3, and antimicrobial filter (18 mL; Medisoft, Sorinne, Begium or 35mL; Air Safety Ltd, Morecambe, UK) according to weight (15-34.9kg, or >35kg, respectively) for which both FRC and cumulative expired volume (CEV) were corrected for (1,17). All children ≥3 years used a mouthpiece interface and nose clip. MBWs were all performed by well-trained operators at each site. All centers followed the same standard operating procedure (SOP) for MBW testing. Quality control was performed by two authors (RMS and PMG) using the same approach for test acceptance (1,17). Exclusion criteria included marked irregular breathing pattern during washout, signs of leak, box-shaped flow-volume loops suggestive of upper airway obstruction due to improper head positioning, or an inappropriately high CO_*2*_-offset during inspiration indicating rebreathing of CO_*2*_ or sensor error.

### Statistical analysis

Characteristics of the study population were assessed using descriptive statistics. All non-parametric outcomes were transformed using the logarithm to base 10 (log_10_), to achieve a more normal distribution of data. In addition, age and height were also log transformed to consider the rapid maturation and growth of the respiratory system during the first years of life, together with a continuously slower maturation and growth towards early adulthood (Appendix Figure 1 a-i). The log-transformed, predefined, clinically relevant, demographic data (log_10_(age), log_10_(height), and log_10_(weight)), together with position (sitting/supine) and gender, were used in multivariate linear regression models for derivation of prediction equations. The residuals for FRC and LCI increased with increasing values (Appendix Figure 1 panels b and e) and for that reason both FRC and LCI were log-transformed (Appendix Figure 1 panels c and f). Prediction equations and z-scores calculations were derived for log_10_(FRC) and log_10_(LCI) (Appendix). The residual standard deviation (RSD) from the linear regression model was used for calculations of z-scores, with the equation: z = (measured value – predicted value)/ RSD. P-values < 0.05 were considered statistically significant. Excel (2010 and 2016) were used for data summaries. Statistica 7 (StatSoft, Tulsa, OK, USA) and SAS Enterprise Guide 7.1 were used for further statistical analyses.

### A new and physiologically reasonably outcome parameter

Whilst log_10_(LCI) was a statistical approach to improve the fitting of data, it did not generate a physiologically reasonable parameter, and alternate approaches in the literature were examined. Arborelius et al.(18) defined the relationship between VD/VT and LCI as: VD/VT = 1 – K/LCI, where K was a constant. The fact that LCI was not linearly related to the actual ratio of dead space ventilation (expressed as volume of dead space per tidal volume, VD/VT)(18) represents an issue for clinical interpretation of longitudinal results as a change in LCI from 7 to 8 is more significant than a change in LCI from 11 to 12(18). The equation by Arborelius et al.(18) can be re-written as VD/VT = 1 - K*1/LCI – showing that a simple inversion of LCI (1/LCI; or calculation as FRC/CEV) should produce a linear relationship with VD/VT and therefore enable comparisons of change across different VI severities. We therefore tested if the relationship shown by Arborelius et al in an adult cohort using N_*2*_MBW could be reproduced in a cohort of children and adults containing a suitably wide range of VI (i.e., including both health and disease). The same linear multivariate regression model used for LCI and log_10_[LCI] was built for derivation of prediction equations for 1/LCI (i.e., FRC/CEV[%]) and it did not require log-transformation for data fitting, as normality assumptions of the residuals were met (Appendix Figure 1 panels g to i). We termed the index “ventilation distribution efficiency; VDE” because an increasing numerical value represents increased gas mixing efficiency in the lungs, which reflects improved ventilation distribution.

### Study population used for physiological validation of “ventilation distribution efficiency”

This physiological validation was performed by examining the relationships between LCI and VD/VT, log_10_(LCI) and VD/VT, and VDE to VD/VT, respectively, across a much wider range of LCI values than was contained within the original healthy cohort. To achieve this, the following cohort was created from a subgroup of the healthy cohort enriched by historical disease cohorts: SF_*6*_MBW data (EXHALYZER D®, Spiroware® 3.2.1) collected from 59 randomly selected healthy Danish and Swedish children aged 0.1 to 3.6 years (median 1.6 years) (as a subgroup from the study cohort); SF_*6*_MBW data from Danish infants and toddler with CF (138 Exhalyzer D® MBWs from 26 patients); and Swedish N_*2*_MBW data from 22 adults with COPD aged 51-71years, and 27 adults with asthma aged 18-63 years.

## RESULTS

### Demographic data of participants for reference equations

Demographic data and MBW outcomes in the 327 children (176 [54%] boys) aged 1 month to 18.4 years, studied on one occasion each were included. The vast majority (n=191 [group 1]; 58%) were under three years of age (22% tested in Denmark, 78% tested in Sweden), and studied in a supine position during quiet sleep (natural sleep n=178 [93.2%] or after Dexmedetomidine sedation n=13 [6.8%]). Median (range) age, weight, and height for group 1 and 2 respectively were 0.90 years (0.14;2.97), 9.8 kg (5.3; 14.9), 75.0 cm (59.0; 97.5) and 8.1 years (3.9;18.4), 28.6 kg (15.5; 81.2), 133.9 cm (99.5; 188.0). As expected, close correlations were seen between age, height, and weight (R^2^>0.95 for all). Mean (SD) BMI was 17.5 (2.29) kg/m^2^ ranging from 12.56 to 27.52kg/m^2^.

Seventeen (11.6%) children above 2 years of age were overweight (using the International Obesity Task Force reference(19)), and six (3.3%) infants had BMI ≥ +2SD according to the 2014 Danish reference population(20). The majority of participants in group 1 (168; 88.0%) provided three technically acceptable trials and 23 (12.0%) provided two trials. All except eight (94.1%) participants in group 2 provided triplicate MBW trials. Mean (SD) CV for the whole dataset of 327 children was 2.5 (1.8)% for FRC, and for LCI it was 2.6 (1.7)%. Median (range) FRC and LCI for group 1 were 179 (71; 414) mL and 6.84 (5.75; 8.21), respectively, and for group 2 was 1065 (452; 3989) mL and 6.03 (5.15; 6.84), respectively.

### Comparisons of z-scores using previously published SF_6_MBW reference equations

The Lum et al study(13) is the only published pediatric SF_6_MBW dataset covering a similar age range to our cohort (2 weeks to 19 years). The prediction equation and FRC z-score calculation given by Lum et al(13) was:

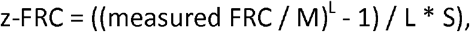

where M = exp (-11.07 + (2.12 * ln height) + (0.27 * age^0.5^) + (0.04 * sex)),

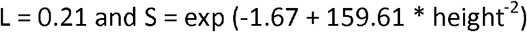

When applied to our dataset it produced a fair fit of z-scores across the age range as a whole: mean (SD) z-score -0.25 (0.91). The strongest agreement (i.e., closest mean z-score values to 0) occurred in school aged children (6-18 years: -0.19 [0.95]) and weakest in younger age groups (0-3 years, -0.41 [0.80]; 3-6 years, +0.48 [1.00]).

The corresponding Lu et al equation derived from a cohort of children aged 0-6 years (14) was: z-FRC = ((measured FRC / M) - 1) / L * S),

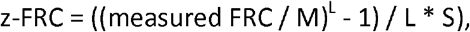

where M = 0.8238 + 0.0740 * age – 0.0068 * age^2^-2.1672 * height + 1.6568 * height^2^ + 0.0075 * sex, L = 0.2597, S = 0.1615

When applied to our dataset, z-FRC values were lower than expected in the age range 0-3 years: mean (SD) -0.81 (0.99), but a good fit in the preschool age range (3-6 years, 0.06 [1.13]) while z-FRC values were markedly different, and greater than expected, in subjects older than 6 years (1.16 [1.81]) with an exponential pattern to this over-estimation with increasing age. It should be noted that this is an older age range than that used in its development (Figure 1A).

**Figure A+B.**
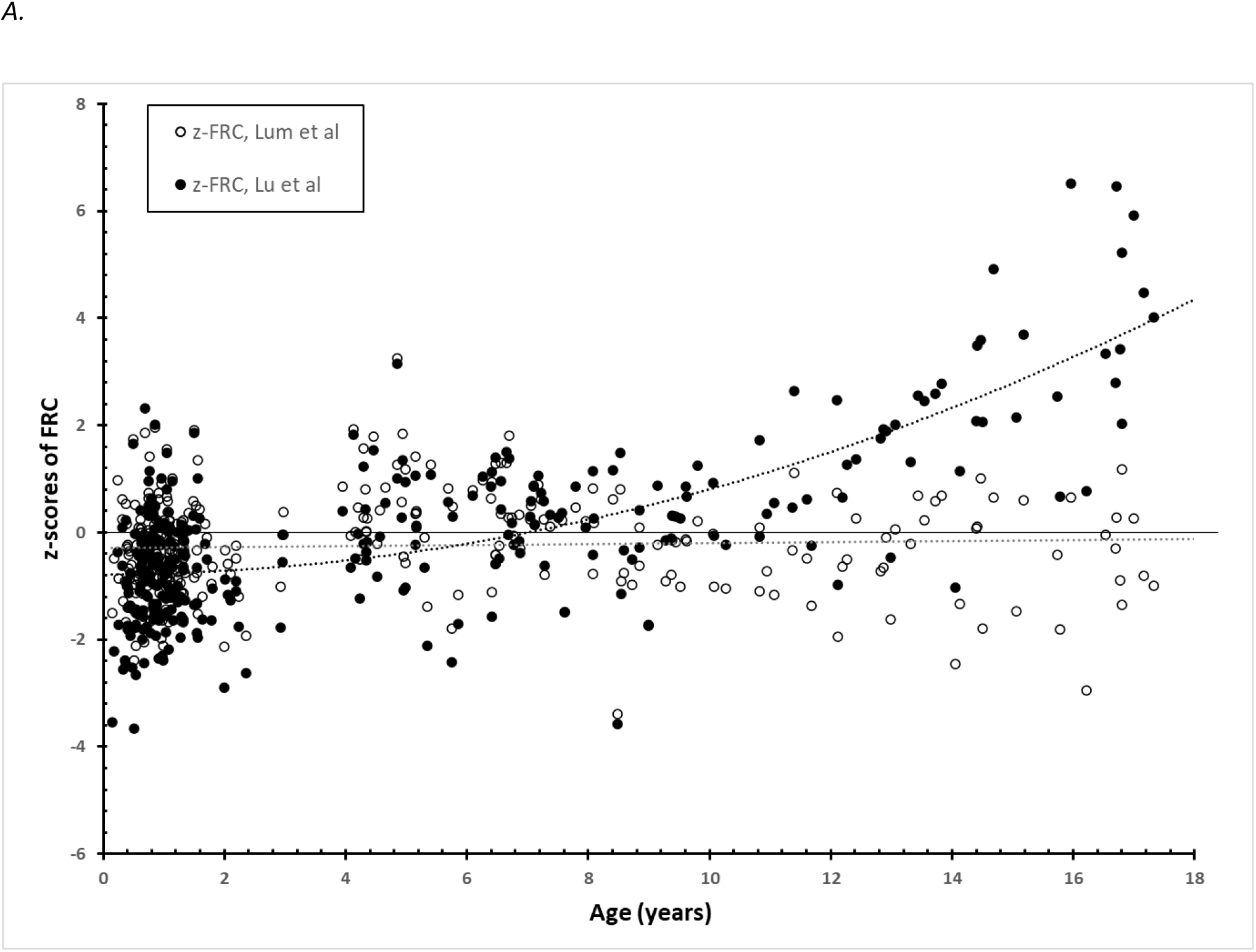
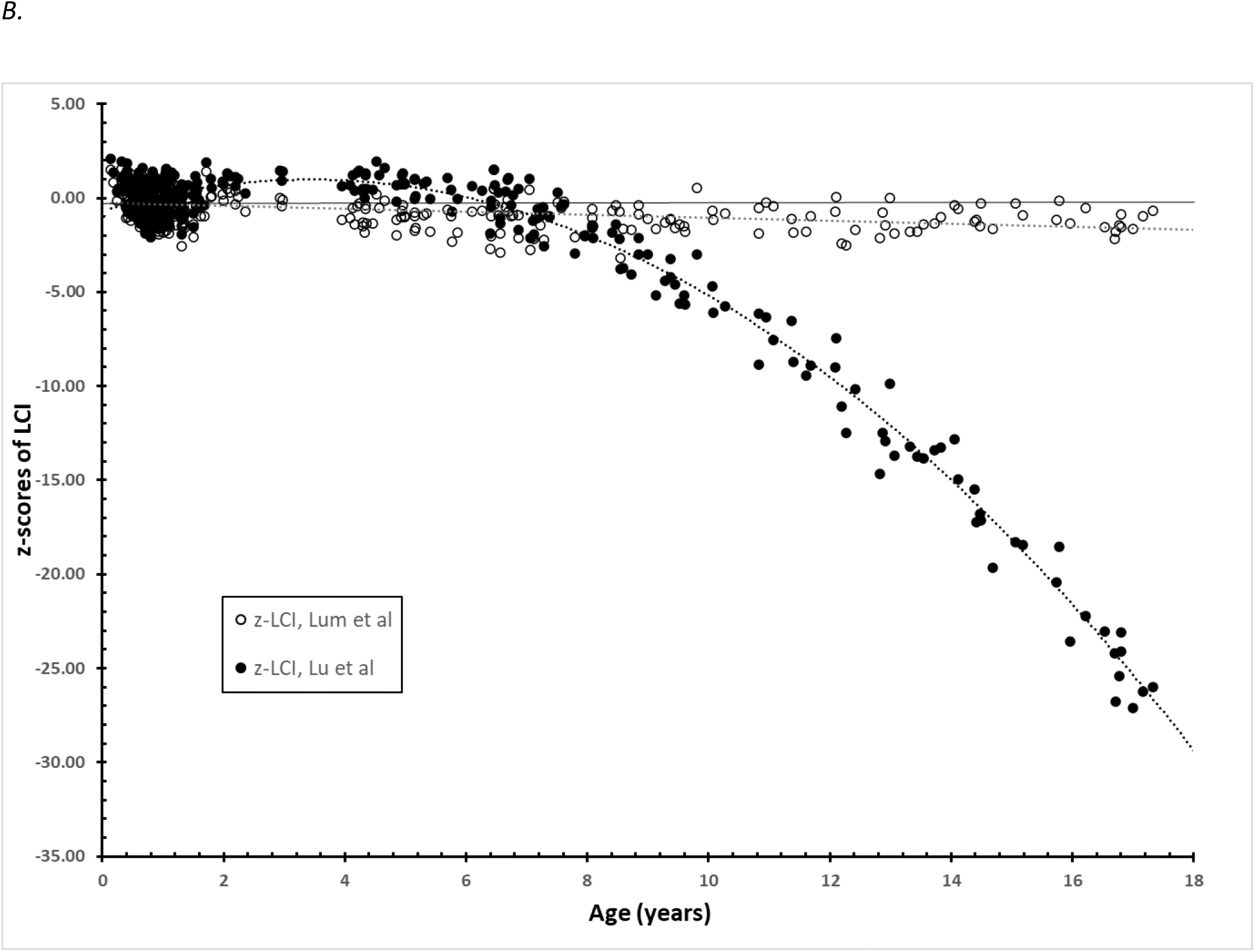
Cohort (327 healthy children aged 0.14 to 18.4 years) z-scores for FRC (panel A) and LCI (panel B) calculated for our cohort using the equations published by Lum et al(13) and Lu et al(14).

For LCI, the prediction equation published by Lum et al(13) was:

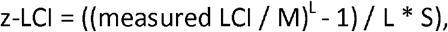

where M = 5.99 + (73.85 * height) ^-1^, L = -0.81, S = 0.08

When applied to our dataset, overall mean z-LCI was lower than expected (mean [SD] -0.61 [0.89]), with the best fit in age 0-3 years (-0.25 [0.81]), and greater deviation encountered in the 3-6 year (-1.03 [0.57]) and 6-18 year age ranges (-1.16 [0.76]).

The corresponding Lu et al(14) LCI prediction equation was:

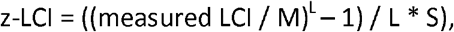

where M = 1.8606 – 1.0399 * age + 0.1210 * age^2^ + 12.5766 * height – 6.3428 * height^2^, L = -0.5565, S = 0.077

Application to our dataset produced a good fit to our data over the first three years of life (0.05 [0.85]), but overestimated z-scores in the 3-6 year old subjects (0.71 [0.57]) and produced extremely poor fit in the 6-18 year old age range (-8.19 [8.39]) (see earlier comments) (Figure 1B).

### SF_6_MBW derived ventilation distribution inhomogeneity and VD/VT ratio

The physiological explanation and calculations first described by Arborelius et al(18) support the notion that ventilation distribution efficiency (VDE) is an index reflecting both anatomic dead space ventilation and ventilation inhomogeneity arising more peripherally in the lungs (see Appendix for further details). On the basis of this argument, a strong linear agreement was found between VD/VT ratio and VDE (i.e. the inverted LCI, FRC/CEV[%]) in our combined set of healthy children (age 0-3.6 years) and children with CF (age 0-3.7 years) (Figure 2). The relationship found in Figure 2 (*VD/VT = 1*.*11 - 0*.*049 * 1/LCI*) was very close to the ideal relationship proposed by Arborelius et al of VD/VT = 1 - K/LCI, where K ≈ 4 (see Appendix) (18). This was also confirmed in a separate dataset of Exhalyzer D® N_2_MBW tests performed in adults with COPD and asthma, producing a very similar regression line (VD/VT = 1.05 -0.044*1/LCI) was found. This justified our approach to pool data from these cohorts to achieve a larger range of ventilation inhomogeneity. We then examined the relationship of VD/VT to LCI and log_10_(LCI) and found a non-linear relationship to both (Appendix Figure 2 panels a and b).

**Figure 2.**
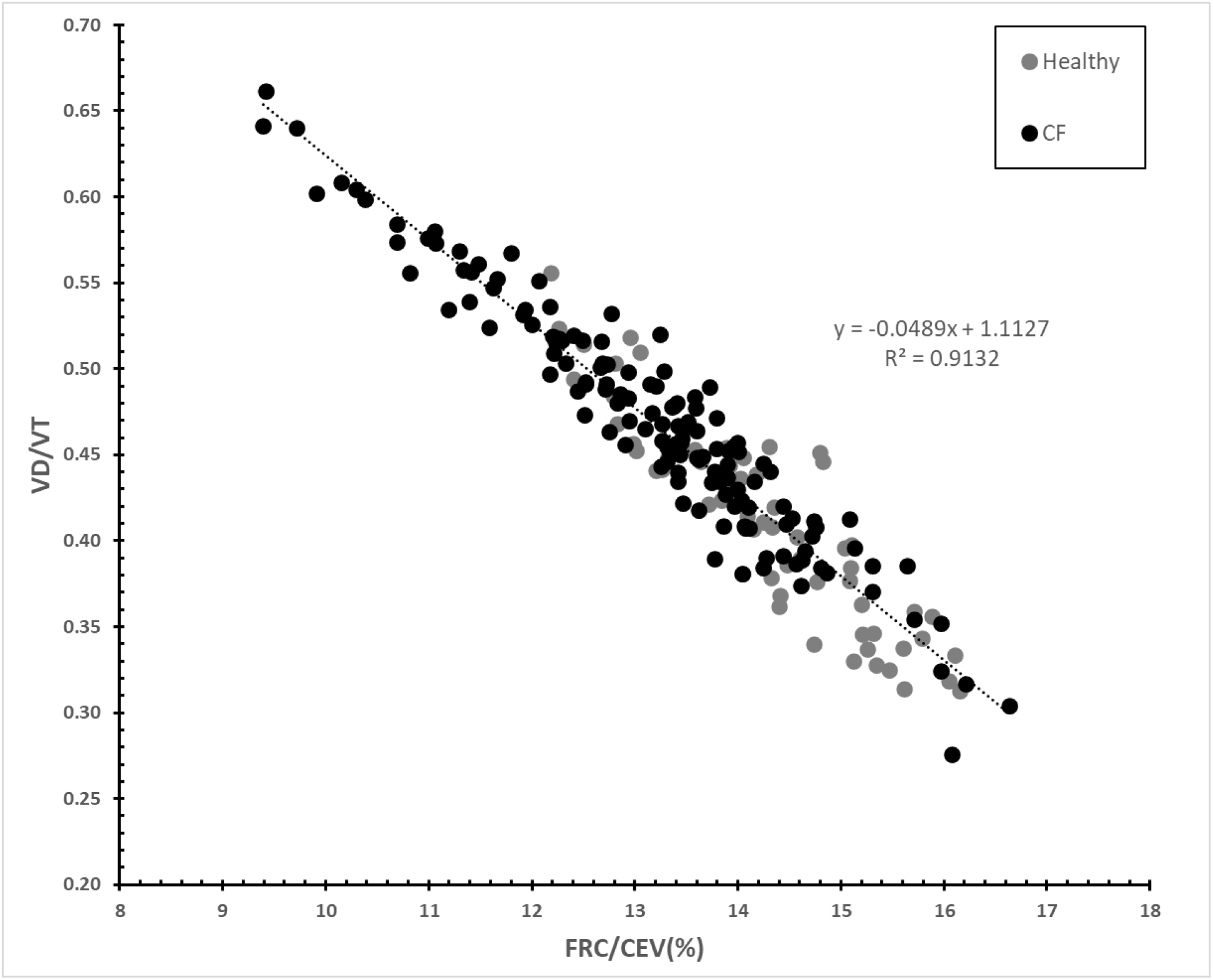
Dead space volume per tidal volume (VD/VT ratio) plotted against ventilation distribution efficiency (VDE; i.e. FRC/CEV[%]) derived from SF_6_MBW in our cohort of 59 healthy children aged 0-3.6 years and 26 healthy children with CF aged 0-3.7 years (total of 138 MBW occasions).

### Reference equations of ventilation distribution efficiency (VDE) and log_10_(FRC)

A multivariate linear regression model was used with VDE as the dependent factor and log_10_(age), log_10_(height) and log_10_(weight), in addition to gender and posture as predictors. Only log_10_(age) (p<0.0001) and gender (p=0.0153) showed statistical significance, with a highly significant overall model fit (R^2^ 0.59, F 237, and p<0.001). Performing a similar multiple regression model with log_10_(FRC) as the dependent variable, found significant predictors to be log_10_(height, cm) (p<0.0001), log_10_(age) (p=0.0339), and posture (p<0.0001), with a highly significant overall model fit (R^2^ 0.98; F 5189; p<0.0001).

### Derived regression equations for VDE and log_10_(FRC)

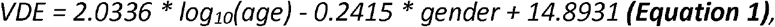

*where age (years), gender (boys = 1 and girls = 0)*.

The 95%CI (confidence interval) was 14.74 to 15.05 (p<0.0001) for intercept, 1.85 to 2.22 for Log_10_(age) (p<0.0001) and -0.44 to -0.05 (p=0.0153) for gender. Residuals were normally distributed with an overall residual standard deviation (RSD) for VDE of 0.8883; varying from 0.8845 in 0-3 years old children, 0.7312 in children age 3-6 years, and 0.9372 in the oldest children (6-18 years); and for boys 0.870 and for girls 0.909.

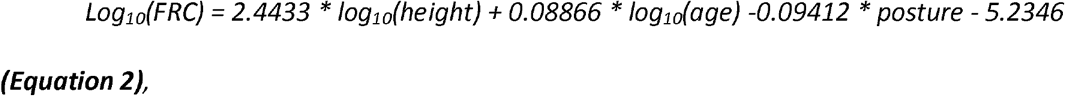

*where height (cm), age (years), posture (supine = 1, upright = 0)*.

The 95%CI was -5.82 to -4.65 (p<0.0001) for intercept, 2.14 to 2.75 for log_10_(height) (p<0.0001), and 0.0068 to 0.17 for log(age) (p=0.0339). Further analysis disclosed a normal distribution of residuals. Overall residual standard deviation (RSD) for log_10_(FRC) was 0.0722, varying from 0.0676 in 0-3 years old children, 0.080 in children age 3-6 years, and 0.0776 in the oldest children (6-18 years).

Z-scores for VDE, and log_10_(FRC) were calculated using the same mean RSD across the whole age range, 0.8883 for VDE, and 0.0722 for log_10_(FRC) (Figure 3 A-B). Mean z-VDE (SD, min; max) was - 0.03 (1.00, -2.65; 2.95) in 0-3 year old children, 0.12 (0.80, -1.70; 1.99) in children age 3-6 years, and 0.01 (1.06, -2.19; 2.95) in the oldest children (6-18 years), respectively.

**Figure 3A.**
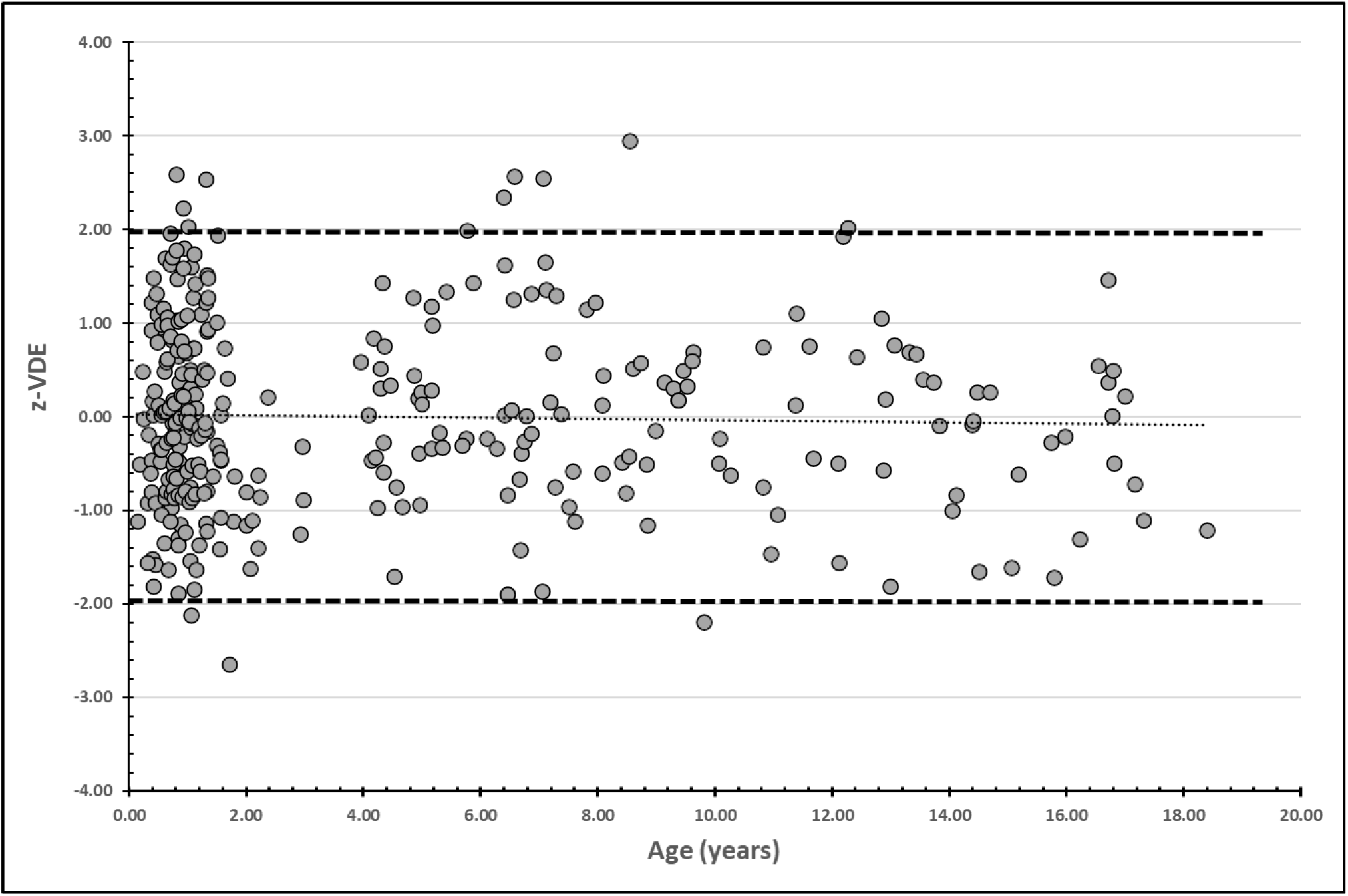
Distribution of SF_6_MBW derived z-scores for ventilation distribution efficiency (VDE; i.e FRC/CEV[%]) vs age in our cohort of 327 healthy children. Upper limit of normal (z-score = +1.96) and lower limit of normal (z-score = -1.96) are given as hatched lines.

**Figure 3B.**
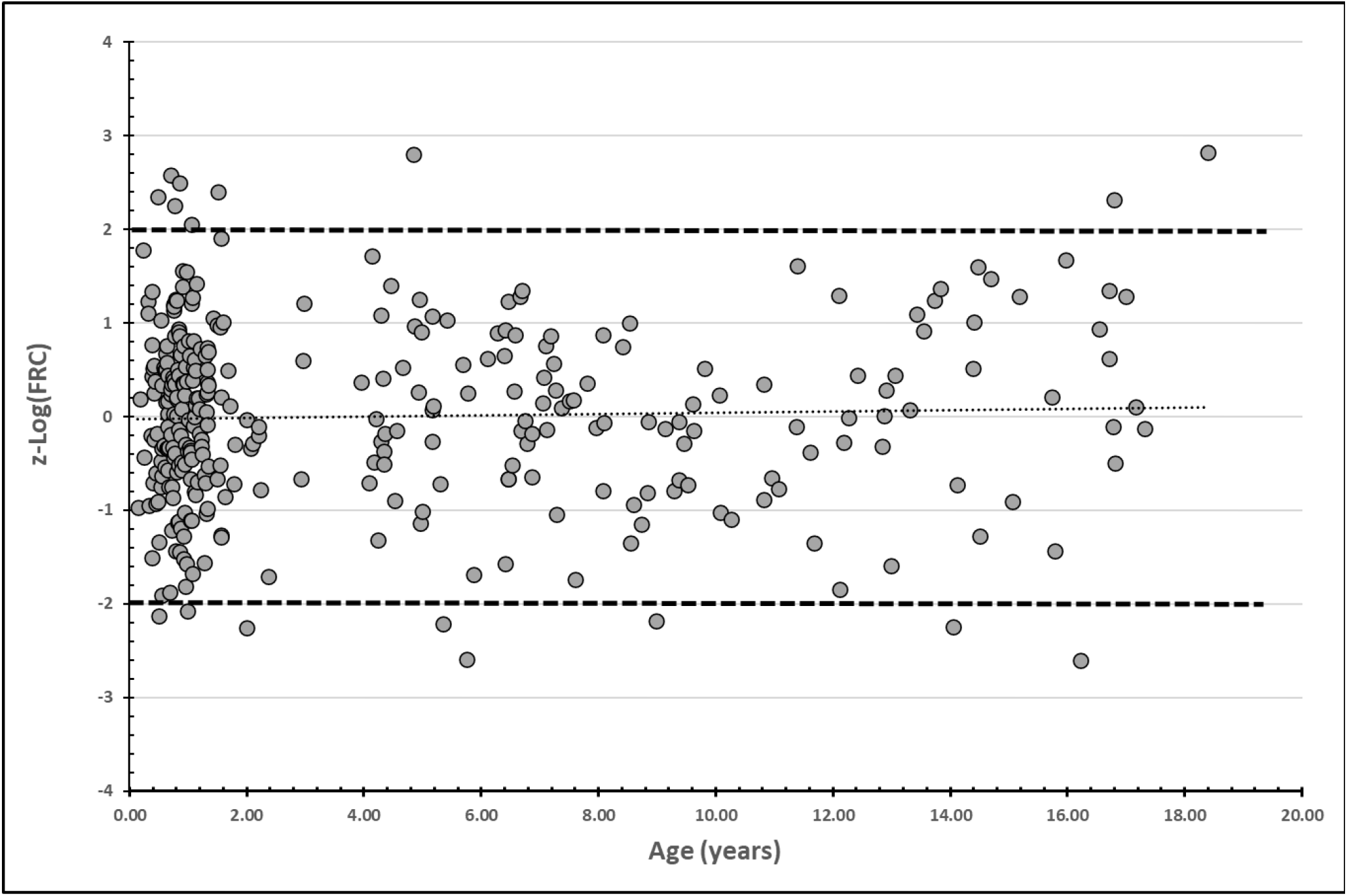
Distribution of SF_6_MBW derived z-scores for log_10_(FRC) vs age in 327 healthy children. Upper limit of normal (z-score = +1.96) and lower limit of normal (z-score = -1.96) are given as hatched lines.

## DISCUSSION

### Summary of findings

This study represents the first and largest SF_6_MBW normative dataset to date using a commercially available device encompassing not only infants and toddlers but the entire pediatric age range. A statistically strong linear regression equation was derived for ventilation distribution efficiency (VDE; defined as FRC/CEV[%]) based on log_10_(age) and gender as predictors. This was physiologically meaningful as these demographic variables are known to be closely associated with the growth and maturation of the respiratory system(9–12) including the gender differences in lung structure seen already during the first years of life(9).

FRC results required log-transformation to achieve a linear relationship with log-transformed height together with log_10_(age) and posture as predictors, also resulting in a strong reference equation. The two resulting regression equations produced residual standard deviations (RSD) that varied minimally across age and gender, allowing for the use of the same RSD across the entire age range in the calculation of z-scores, which also showed an even distribution across age (Figure 3 A and B).

While LCI is currently the most widely reported global ventilation distribution outcome in paediatric studies, the fact that LCI was not linearly related to actual ratio of dead space ventilation (VD/VT) in our dataset makes it not ideal for clinical interpretation or follow-up studies. A change in LCI from 7 to 8 reflects a more significant change in VI than that when LCI changes from 11 to 12 (see further explanation in Appendix). This is not a new issue: increasing variability with increasing LCI has been reported, and adjusted for in variability studies by log-transformation of LCI data (log_10_[LCI]), thus advocating to treat changes in LCI as a percentage change, when back transforming data(21). Several other studies have also used percentage LCI change as a research outcome(22–24). However, as demonstrated in the results of this present study, a simple inversion of LCI (ventilation distribution efficiency; VDE) resulted in a linear relationship with VD/VT and facilitated comparisons of outcomes across severities, which LCI, log_10_(LCI) and its z-scores did not (see Appendix).

There are important differences between the two published SF_6_ reference studies to date. The multi-center study by Lum et al included 497 healthy children aged 2 weeks to 19 years, tested on 659 occasions(13), while the Lu et al was a single-center study involved 280 healthy infants and children up to age 6 years, tested on 644 occasions(14). Gender and log-transformation of age were not used as predictors in those two previous SF_6_MBW LCI studies(13,14), in contrast to our study. Including gender for prediction of VI is highly relevant as the larger number of alveoli in the male vs. female lungs(9) should result in greater structural asymmetries within a given lung volume – and therefore also greater VI. Lum et al used inverted height(13) and Lu et al used a second-degree polynomial of height and age(14) for LCI prediction. The application of a second order polynomial equation explains the highly skewed z-scores for children above age 6 years when applied to our dataset, as it resulted in a mathematical parabola (U-formed curve). Z-scores for children above age 6 years are presented in Figure 2 to reinforce the importance of only applying a reference equation to a study population of the same age range as that developed from. This is in keeping with the authors’ statement, that their newly developed reference equations provide normative data for cohorts with similar population characteristics that utilize the same equipment (RMS)(14).

Both previous LCI studies(13,14) employed a generalized additive model of location, shape and scale (GAMLSS) regression method, by which the skewness of the response variable is modelled by L (lambda), the median by M, and the coefficient of variation is modelled by S(25), as also used in GLI spirometry reference equations(26). Despite this approach, the interpretation of data becomes less intuitive as z-scores derived were not linearly related to absolute LCI values, as illustrated when simulating different LCI values for subjects with a specific height and age (Appendix Figure 3 a + b). Using the Lu et al reference equation (13), an increase of LCI from 6.0 to 8.0 in a 6-month old infant with a body length of 65 cm corresponds to an increase of LCI z-score from -2.69 to +2.72 (delta 5.43), while an LCI increase from 12.0 to 14.0 corresponds to an increase of z-score from +8.49 to +10.23 (delta 1.74) (Appendix Figure 3a). The same pattern of decreasing z-score change for a given actual change in LCI occuring at a higher baseline LCI value was observed with the Lum et al reference equation (Appendix Figure 3b), and also using z-scores of log_10_(LCI) (Appendix Figure 3c) in cases of LCI values above 10. This simulation demonstrated a non-linear relationship between LCI and calculated LCI z-scores, while a linear relationship remained between VDE z-scores and absolute VDE (Appendix Figure 3d). A linear relationship was also found between z-log_10_(LCI) and log_10_LCI values. However, there are two important issues to acknowledge with log_10_(LCI) as an outcome parameter: i) the log_10_(LCI) is not linearly related to VD/VT, and ii) the values of log_10_(LCI) are not physiologically intuitive for clinical interpretation.

We believe that z-scores should be linearly related to the physiological variable it reflects, and as such that changes in z-scores should directly translate in a linear manner to changes in disease severity. We argue that the new index - efficiency of ventilatory distribution (VDE; FRC/CEV(%)) - fulfills such criteria, and therefore should be a preferred MBW index of global ventilation inhomogeneity moving forward.

### Further arguments for VDE as an alternative to LCI

The FEV_1_/FVC ratio (forced expiratory volume in one second / forced vital capacity) is the well-known spirometry index of airway obstruction. The FEV_1_/FVC ratio describes how efficiently the entire volume of the lungs are emptied during a maximal forced expiration of 1 second. Describing ventilatory efficiency in a similar fashion for the MBW method would equate to FRC/CEV, i.e. how efficiently is FRC cleared of an inert marker gas. The larger the CEV required for clearing a given FRC, the less efficient the respiratory system. An LCI of 6.0 will then correspond to an FRC/CEV ratio of 0.167, or 16.7%, i.e. 16.7% of FRC is cleared per one FRC volume of air expired during the washout phase, and an LCI of 12.0 will correspond to a reduced clearing efficiency of 8.3%. The larger the CEV required for a given FRC to clear the tracer gas, the less efficient the respiratory system performance.

The regression equation (Equation 1) gives an VDE increase of 2.03 percentage-points for every log unit increase of 1 in log_10_(age): the predicted VDE increases from 12.9% at age 0.1 year to 14.9% at age 1.0 year, from 14.9% to 16.9% from age 1.0 to 10.0 years. The change in VDE through adolescent years was minimal (17.4% at age 18 years). Whilst we would expect VDE to increase by 2.03% from 10 to 100 years of age, we did not study the lung function in adults within this dataset and remains to be validated.

A one percentage-point reduction in VDE was found to be linearly related to an increase of dead space ventilation (VD/VT) by 4.9 percentage-points (Figure 2A), irrespective of overall ventilation efficiency severity. Importantly, this relationship between VDE and VD/VT was almost identical in the cohort of healthy young children or children with CF performing SF_6_MBW (Figure 2A), to that in adults with obstructive lung disease. The close findings to the Arborelius et al study(18) and these three cohorts reported (children, healthy and with CF, and adults with lung disease) demonstrate the robustness and physiological relevance of the relationship between VDE and VD/VT. In contrast, the relationship between the functional VD/VT ratio and LCI was curvilinear in children with CF, healthy children, and adults (Appendix Figure 2a), and as originally demonstrated by Arborelius et al(18). Also, z-scores for VDE vs age at simulated fixed values for LCI and VDE respectively, demonstrated equidistant z-scores vs. VDE but not vs. LCI over the entire age range from 0 to 18 years (Appendix Figure 3 a+b). A lung function variable should relate linearly to the severity of the condition it aims to reflect, which LCI (CEV/FRC) did not.

### Impact of replacing LCI with ventilation distribution efficiency (VDE)

The problem of non-linearity with LCI and VD/VT might result in problematic evaluations of treatment effects and changes in disease severity over time in previous studies, as an improvement in LCI from 15 to 13 is not as great an improvement of dead space (ventilation distribution inhomogeneity) as a change from 9 to 7. Log-transformation of data (log_10_[LCI]) for the analysis, would partly solve the problem by giving percentage changes over time. As seen in Appendix Figure 2b, the log-transformation only solve the problem of non-linearity in patients with LCI less than 10. Studies using changes in LCI as endpoints might therefore have resulted in exaggerated conclusions.

However, using VDE and the z-VDE instead of LCI or even log_10_(LCI), the changes over time would become smaller in absolute values – especially in more diseased children, but the variation between results would also be less, why the confidence intervals would be narrower and results likely still be significant. This was seen in the study by Robinson et al.(27) after correcting LCI data with the new improved Spiroware® 3.3.1 software, that caused values of LCI to decrease, but at the same time reduced between-subject variation. This resulted in narrower confidence intervals and better statistical significance levels(27). Future studies correcting LCI to VDE, will be needed to fully understand the impact of inverting LCI.

### Strength and limitations

The present study provides robust SF_6_MBW derived linear regression equations for ventilation distribution efficiency (VDE, i.e. 1/LCI or FRC/CEV[%]) and log-transformed lung volume (FRC) covering the whole pediatric age range. It is only the second SF_6_MBW study of normative values to encompass the entire pediatric age range, but the only study based on a commercially available system. The majority of study participants were under the age of 3 years, which is both a strength and a limitation. The large numbers of infants and toddlers ensures detailed description of change during rapid lung maturation and growth. However, the very small number of participants in the 2-4 years age range is a weakness and is a technically challenging period to test either supine during sleep or upright sitting (awake). However, we do not believe, that the gap in the dataset seriously affects the integrity of the reference equations, as a very nice-fitting trendline can be drawn between data of participants 0-2 years andC those 4-18 years (Appendix Figure 1a,d,g). The data used are sufficient to outline the strong physiological arguments for using ventilation distribution efficiency (VDE; FRC/CEV[%]) and z-VDE as a new and superior outcome measure for global ventilation distribution inhomogeneity.

### Summary and conclusion

The previous reference equations acquired using the respiratory mass-spectrometer were not applicable to SF_6_MBW data derived using the Exhalyzer D® over the entire pediatric age range. The physiological rationale for using ventilation distribution efficiency (VDE) as an alternative to LCI in the analysis and interpretation of MBW recordings is presented. VDE was linearly related to the functional respiratory VD/VT ratio and therefore to disease severity. Robust and physiologically relevant reference equations for Exhalyzer D® SF_6_MBW based ventilation distribution efficiency and lung volume are therefore presented for infancy to late adolescence. Derived z-scores of VDE were directly proportional to VDE strengthening its case for future use.

## Data Availability

All data produced in the present study are available upon reasonable request to the authors

## Acknowledgement

The authors are very grateful to all the children and their parents for their participation, and to the staff at the Department of Pediatrics, General Hospital, Skövde, Sweden, the staff at the Gothenburg CF Center, Queen Silvia Children’s Hospital, Gothenburg, Sweden and at the Paediatric Pulmonary Service of Copenhagen University Hospital for their assistance.

## Conflict of interest statement

None of the authors have or have had any financial relationships with ECO MEDICS AG or any organizations that might have an interest in the submitted work. ECO MEDICS AG are not involved in any way in this study. However, Per Gustafsson has spent a large part of his research career on developing and improving the multiple breath washout technique. He has never had any economical relationships with ECO MEDICS AG but has become a close collaborator of the company as he has provided knowledge and respiratory mass-spectrometer for the ECO MEDICS AG for them to be able to perform the gas calibrations leading to the correction equations. The main interest for Per Gustafsson has only been to improve the method as it is needed for the patients.

## APPENDIX

### Mass spectrometer vs. Exhalyzer D® - technical aspects

Differences in technical design are an important factor when comparing reference data. The previously published reference data(13,14) were performed using SF_6_MBW with a fast-responding respiratory mass spectrometer (RMS) based equipment system, which is commonly viewed as the gold standard method. However, this method is not commercially available. A few years ago, our group described novel methodology for SF_6_MBW using this commercially available device (EXHALYZER® D; ECO MEDICS AG, Duernten, Switzerland) and demonstrated excellent agreement between Exhalyser®D and RMS based SF_6_ signals(16). An optimized synchronization approach (“dynamic delay correction”) of gas signals in relation to respiratory flow was applied in the EXHALYZER D® software(16) and is now used as standard within this equipment. Such an option was however not available in the RMS software (developed by one of the current authors, PG) at the time of the reference equation publications by Lum et al(13) and Lu et al(14), which may have contributed to differences in results. Furthermore, a pneumotachometer was used for respiratory flow in the RMS-based MBW studies (13,14), while the EXHALYZER D® device included an ultrasonic flowmeter, which is not sensitive to the dynamic viscosity of the gas measured, i.e. gas composition.

### Statistical Methods

The scatter plots that justified log-transformation of data to achieve linear relationships between lung function outcomes and age and height are shown below (Appendix Figure 1 a-i).

### Derivation of reference equation for log_10_(LCI)

The same analysis used for the derivation of reference equations for LCI and VDE, was performed for Log_10_(LCI). Log_10_(age) (p<0.0001) and gender (p=0.006) showed significant contributions

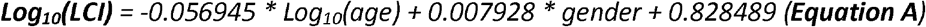

*where age (years), gender (boys = 1 and girls = 0)*.

95%CI (confidence interval) for intercept was 0.823936 to 0.833043 (p<0.0001), -0.062295 to - 0.0515955 for Log_10_(age) (p<0.0001) and 0.002325 to 0.013532 (p=0.006) for gender. Overall model fit was R^2^ 0.56; F 211; p<0.0001. Further analysis disclosed a normal distribution of residuals. Overall residual standard deviation (RSD) for log10(LCI) was 0.02504; for boys 0.02457 and for girls 0.02556.

### VD/VT in relationship to LCI and log_10_(LCI)

Arborelius et al (18) presented a method to calculate the ratio of the functional purely respiratory dead space per tidal volume (VD/VT) from N2MBW recordings, i.e. ventilation to perfusion ratio not included. This functional dead space includes both the airway dead space (series dead space) and the remaining “alveolar” dead space (i.e. the parallel dead space), and is calculated in analogy with the physiological dead space(28): VD/VT = 1-F_E_N_2_ / F_i*d*_N_2_, where F_*E*_N_2_ represents the average end-tidal fraction of N_2_ over a washout, and F_*id*_N_2_ represents the ideal fraction of N_2_ over a washout with the same number of breaths. For this, the dilution factor (W) of the washout is calculated:

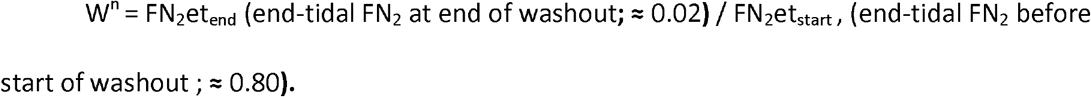

Given these numbers 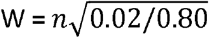. Then, F_*id*_ N_2_ = (0.80 x W + 0.80 x W^2^+ + 0.80 x W ^n^)/n.

Most importantly, the Arborelius et al(18) demonstrated that LCI was non-linearly related to VD/VT:

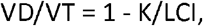

where K = (FN_2_et_*start*_-FN_2_et*end*)/FidN_2_, and typically K = (0.80-0.02)/0.19 = 4.105, and where K represents the optimal (minimal) LCI achievable for a given number of breaths, and the number of breaths is used to determine FidN_2_.

The equation demonstrates that VD/VT is linearly related to 1/LCI (VD/VT = 1 – K*1/LCI), which is therefore a linear representation of the ventilation distribution efficiency (VDE), an index reflecting both anatomic dead space ventilation and ventilation inhomogeneity arising more peripherally in the lungs.

We tested this in N_*2*_MBW data from an adult cohort including healthy elderly subjects and patients with asthma or COPD and reproduced these relationships (data not shown) and found the relationships to be almost identical with those of SF_*6*_MBW data from both healthy children and children with CF. For this reason, they were combined and plotted together (Appendix Figure 2 a-b).

Appendix Figure 2a demonstrates that a change in LCI from 6.0 to 8.0 has greater physiological implications than a change from 14.0 to 16.0, as the change in VD/VT increases more in the lower ranges of LCI changes. This also explain why several previous MBW research publications have reported increasing variation in LCI with higher values. This problem was solved by log-transformation of LCI data – and thus a percentage change, when back transforming data (21,23). However, log_10_(LCI) did not have a linear relationship to VD/VT – especially in the range of LCI above 10 (above log_10_[LCI]=1), (Appendix Figure 2b). This also explains why despite the approach of using percentage change for LCI, the utility of LCI in children with severe lung disease appears limited.

### Z-scores vs absolute values for LCI, log_10_(LCI) and VDE, respectively, in simulation using equations from previous studies(13,14) and the current study

Simulating fixed ages and heights with different values of LCI and VDE, we found a curvilinear relationship between z-LCI and LCI using both of the two previously published reference equations(13,14) (Appendix Figure 3a and 3b). Using Equation A from the current study z-log_10_(LCI) and log_10_(LCI) were linearly related, but z-log_10_(LCI) was not linear related to LCI (Appendix Figure 3c). Using Equation 1 from the current study z-VDE and VDE were linearly related (Appendix Figure 3d). Using a log approach to LCI for clinical interpretation is likely to be challenging. We argue that VDE – ventilation distribution efficiency – is a better option as it is physiologically meaningful, linearly related to VD/VT and easier to understand and interpretate.

### Z-scores for VDE vs age and z-scores for log_10_(LCI) vs age at simulated fixed values for LCI and VDE respectively, demonstrating equidistance for z-scores in relation to VDE values but not in relation to LCI values

We simulated LCI values of 7, 8, 9, 10, 11, 12 and 13 in subjects aged 0.1-18.4 years. From these data, we calculated the z-scores of VDE (Appendix Figure 4a) and z-log_10_(LCI) (Appendix Figure 4b). For a given LCI value the corresponding z-VDE and z-log_10_(LCI) both indicated worse lung function with increasing age, i.e LCI decreases with age in healthy children, as seen in our data and also in two previous publications(13,14). Differences in z-VDE and in z-log_10_(LCI) between two given LCI-values (the space between the lines in Appendix Figure 4 a+b) decreased with severity of LCI - identically across all ages.

Secondly, we simulated VDE values of 17%, 16%, 15%, 14%, 13%, 12%, and 11% across all ages for z-VDE (Appendix Figure 4c) and for z-log_10_(LCI) (Appendix Figure 4d). Here we found the changes between z-VDE to be exactly the same for all differences in VDE values (same distance between the lines in Appendix Figure 4c), but this was not the case for z-log_10_(LCI) (Appendix Figure 4d). This shows that the relationship of z-VDE is linearly related to the efficiency of ventilatory distribution (VDE) – and therefore also to VD/VT.

#### Comparison of z-log_10_(LCI) and z-VDE

To enable comparison of the calculated z-scores of Equation 1 (z-VDE) and Equation A (z-log_10_[LCI]) we first multiplied the z-log_10_(LCI) by minus 1 to ensure the same direction for severity of disease – that is, smaller z-scores meaning greater disease. From our cohort of healthy children and children with CF, we found that within the normal range of +2 to -2 z-scores, the difference increased towards both ends of the interval. However, looking at the differences between the two z-scores in children with CF, we found the difference to increase with greater lung involvement (Appendix Figure 5). We believe this reflects the fact the log_10_(LCI) does not completely compensate for the non-linearity to VD/VT in cases of more ventilation inhomogeneity.

**Appenix Figure a-i.**
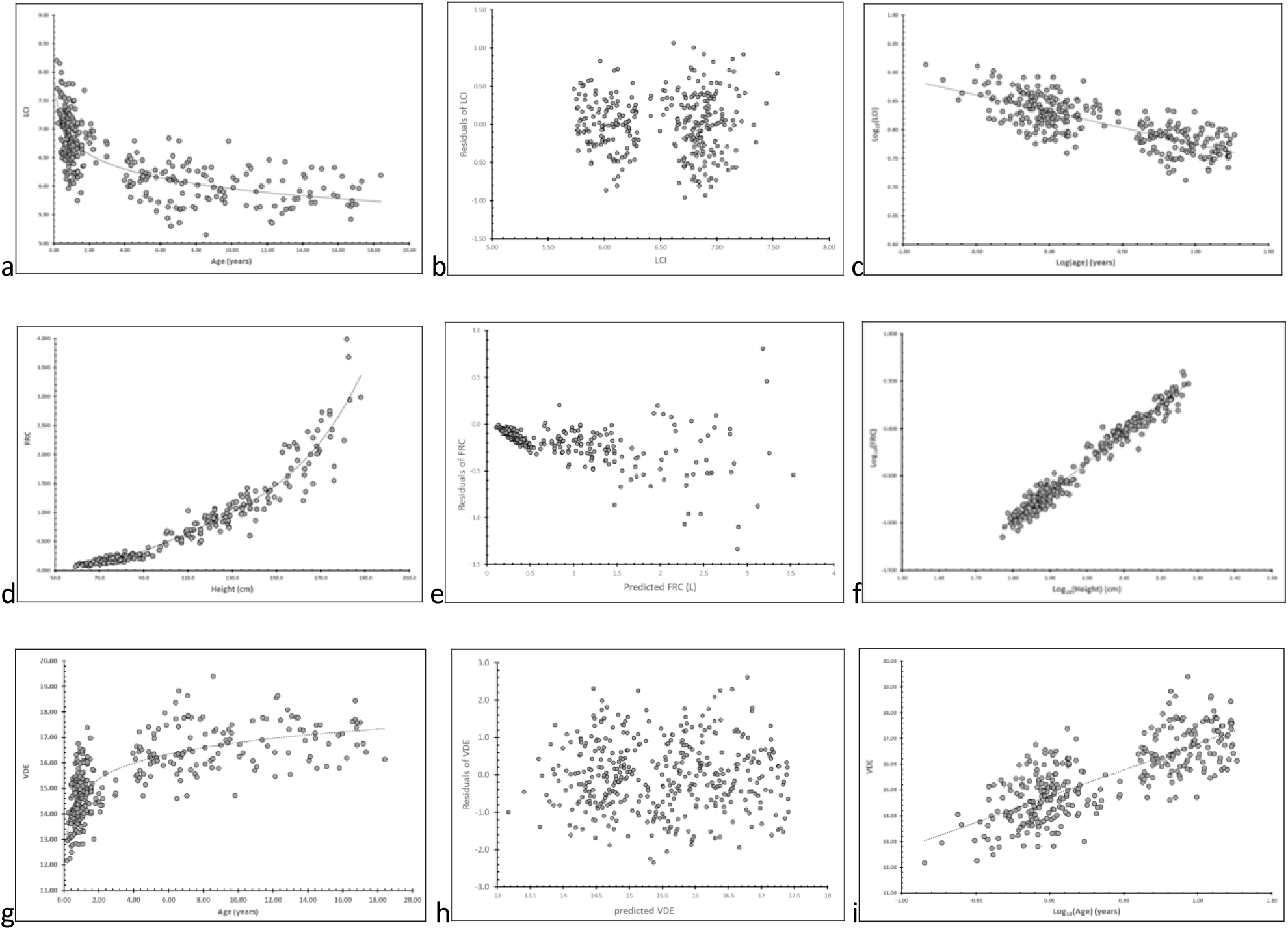
SF_6_ MBW outcomes from 327 healthy children aged 0.14 to 18.4 years in relation to age, log_10_(age), height and log_10_(height), respectively.

**Appendix Figure 2a.**
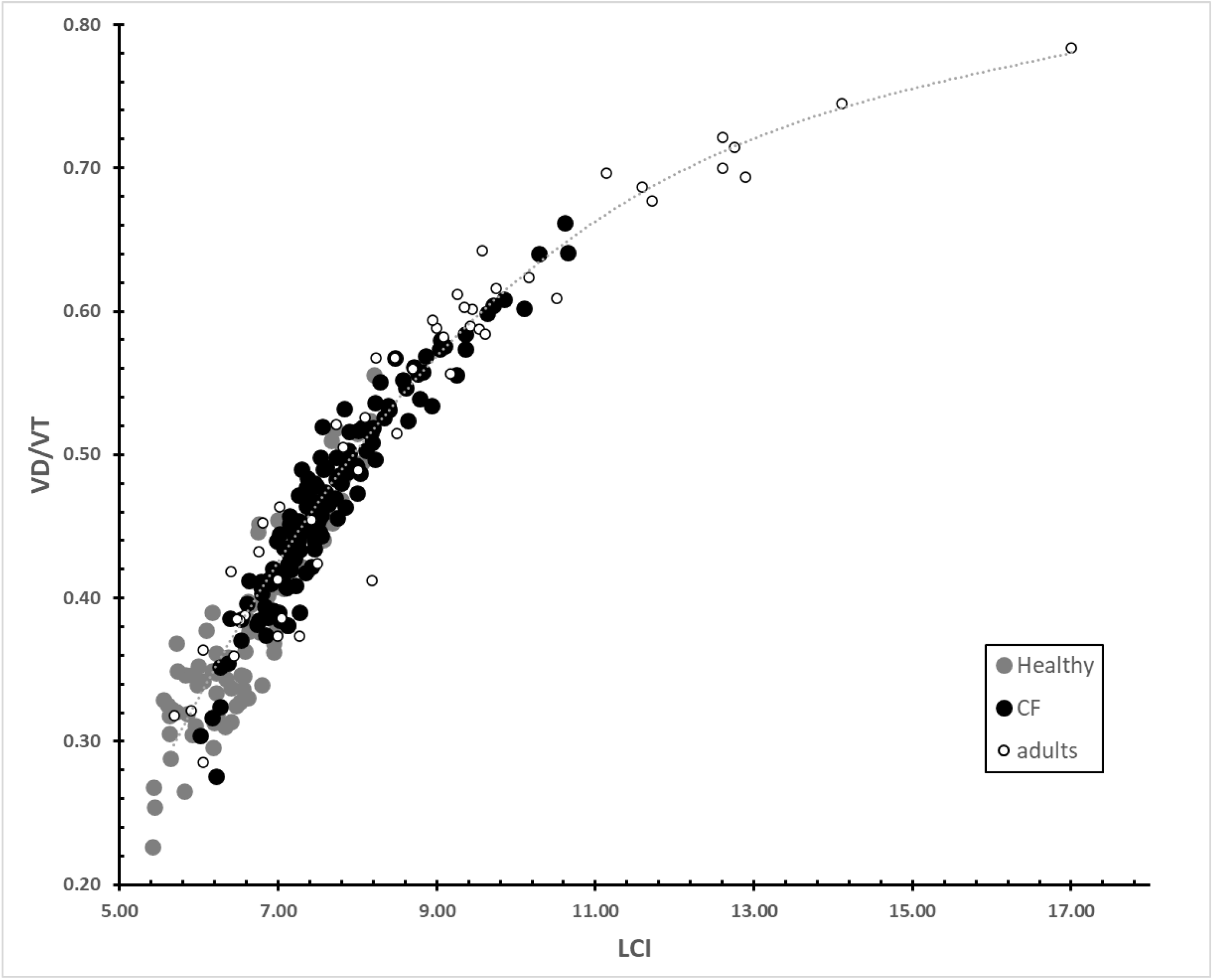
Dead space volume per tidal volume (VD/VT ratio) plotted against the LCI derived from SF_6_MBW in 59 healthy children aged 0-3.6 years, 26 healthy children with CF aged 0-3.7 years (138 MBW sessions), and from N_2_MBW in 22 subjects with COPD aged 51-71years, and 27 subjects with asthma aged 18-63 years, giving a non-linear relationship.

**Appendix Figure 2b.**
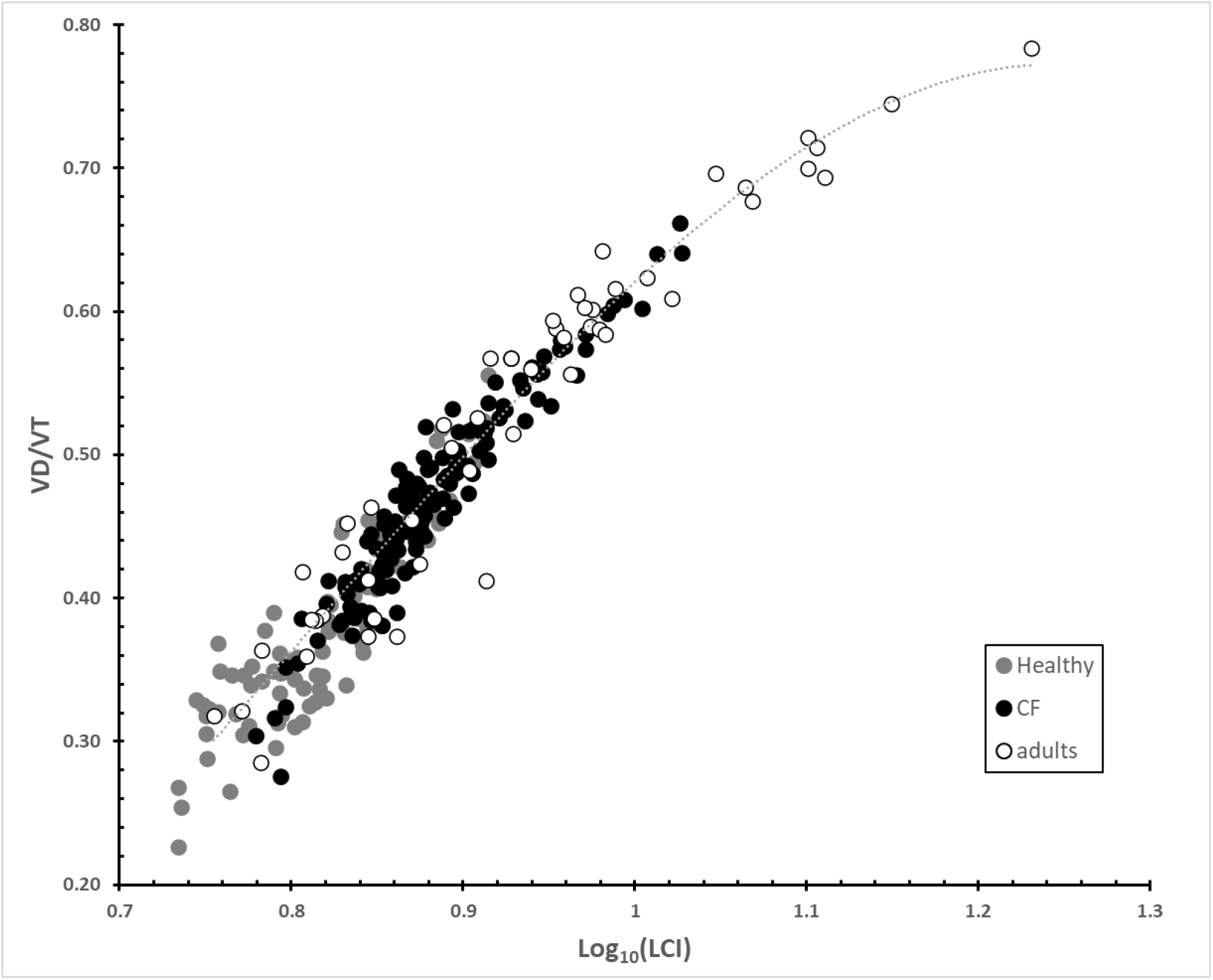
Dead space volume per tidal volume (VD/VT ratio) plotted against the log-transformed lung clearance index (log_10_[LCI]) derived from SF_6_MBW in 59 healthy children aged 0-3.6 years, 26 healthy children with CF aged 0-3.7 years (138 MBW sessions), and from N_2_MBW in 22 subjects with COPD aged 51-71years, and 27 subjects with asthma aged 18-63 years, giving a non-linear relationship.

**Appendix Figure 3.**
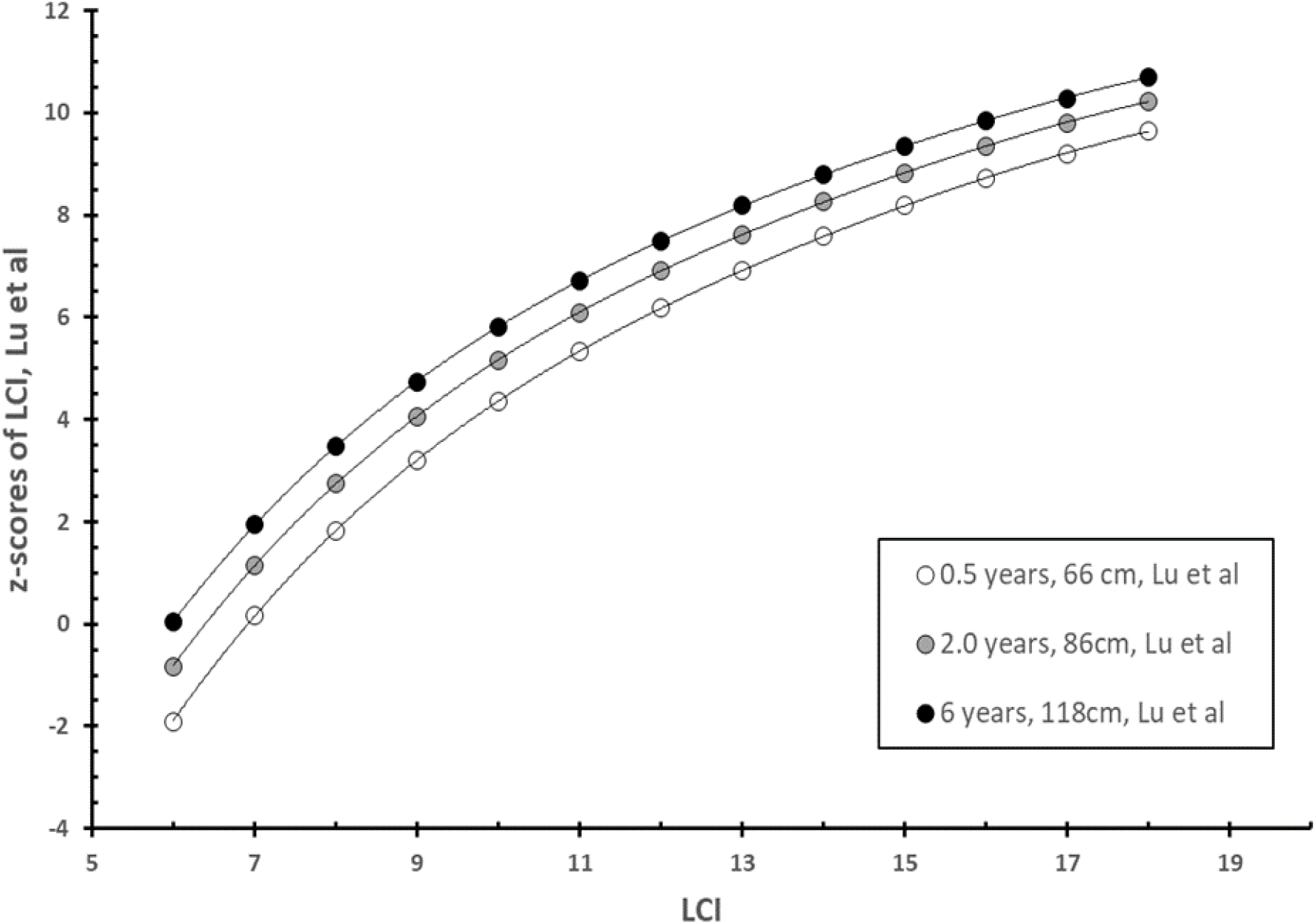
LCI z-scores vs LCI with three different simulated values for heights and ages using the Lu et al reference equation (13)

**Appendix Figure 3b.**
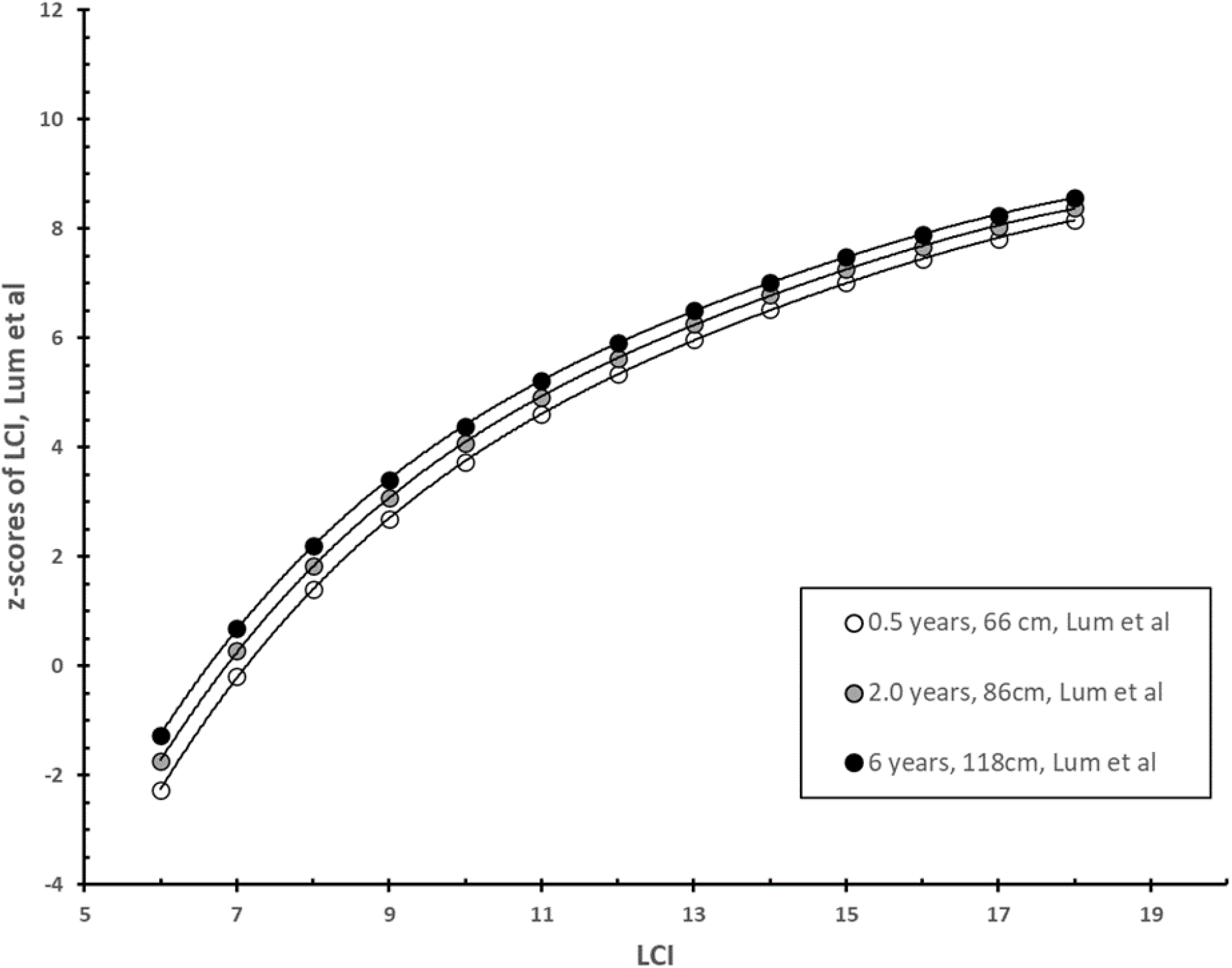
LCI z-scores vs LCI with three different simulated values for heights and ages using the Lum et al reference equation (12)

**Appendix Figure 3c.**
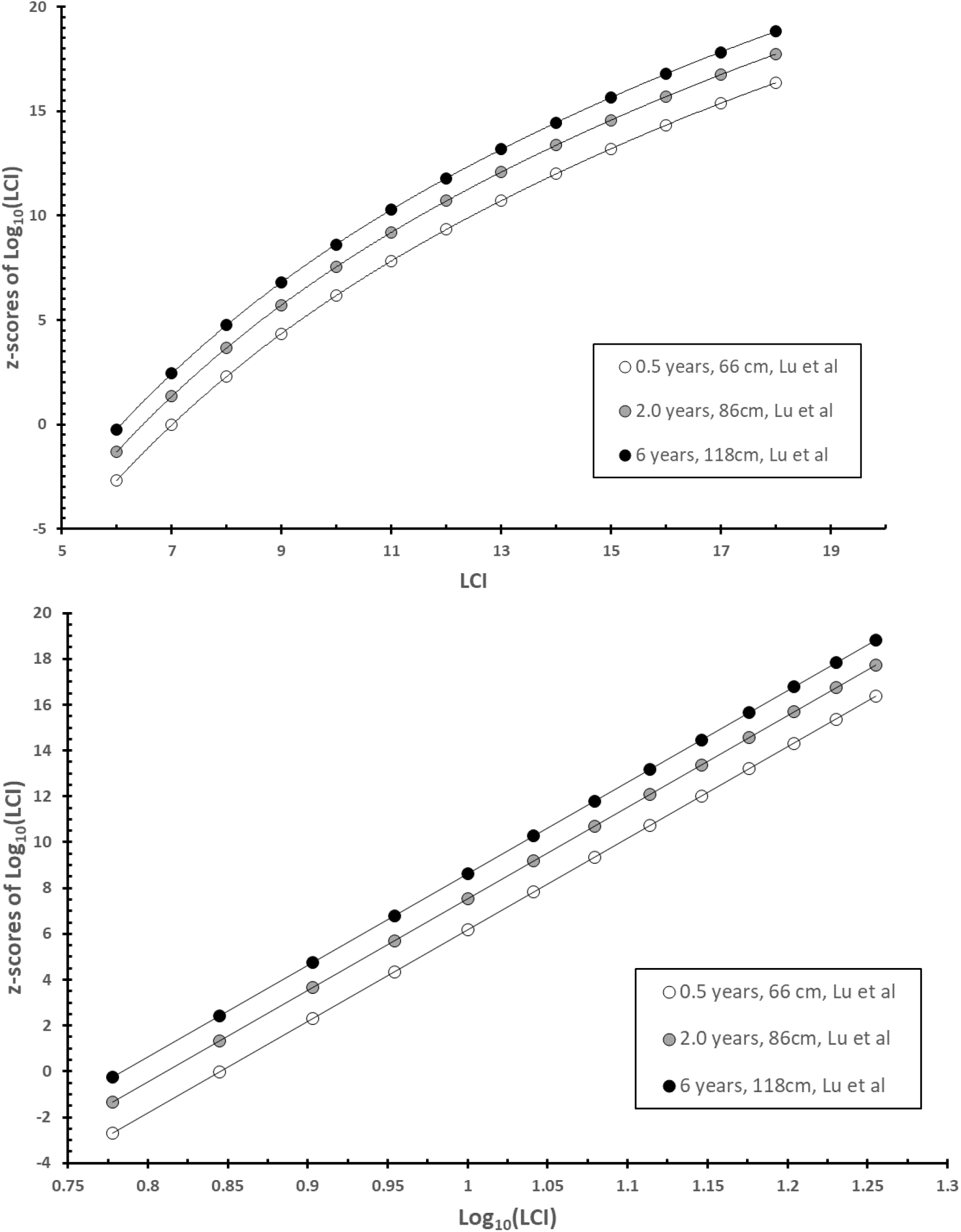
Log_10_(LCI) z-scores vs LCI and log_10_(LCI) with three different simulated values for heights and ages using Equation A of the Appendix in this paper

**Appendix Figure 3d.**
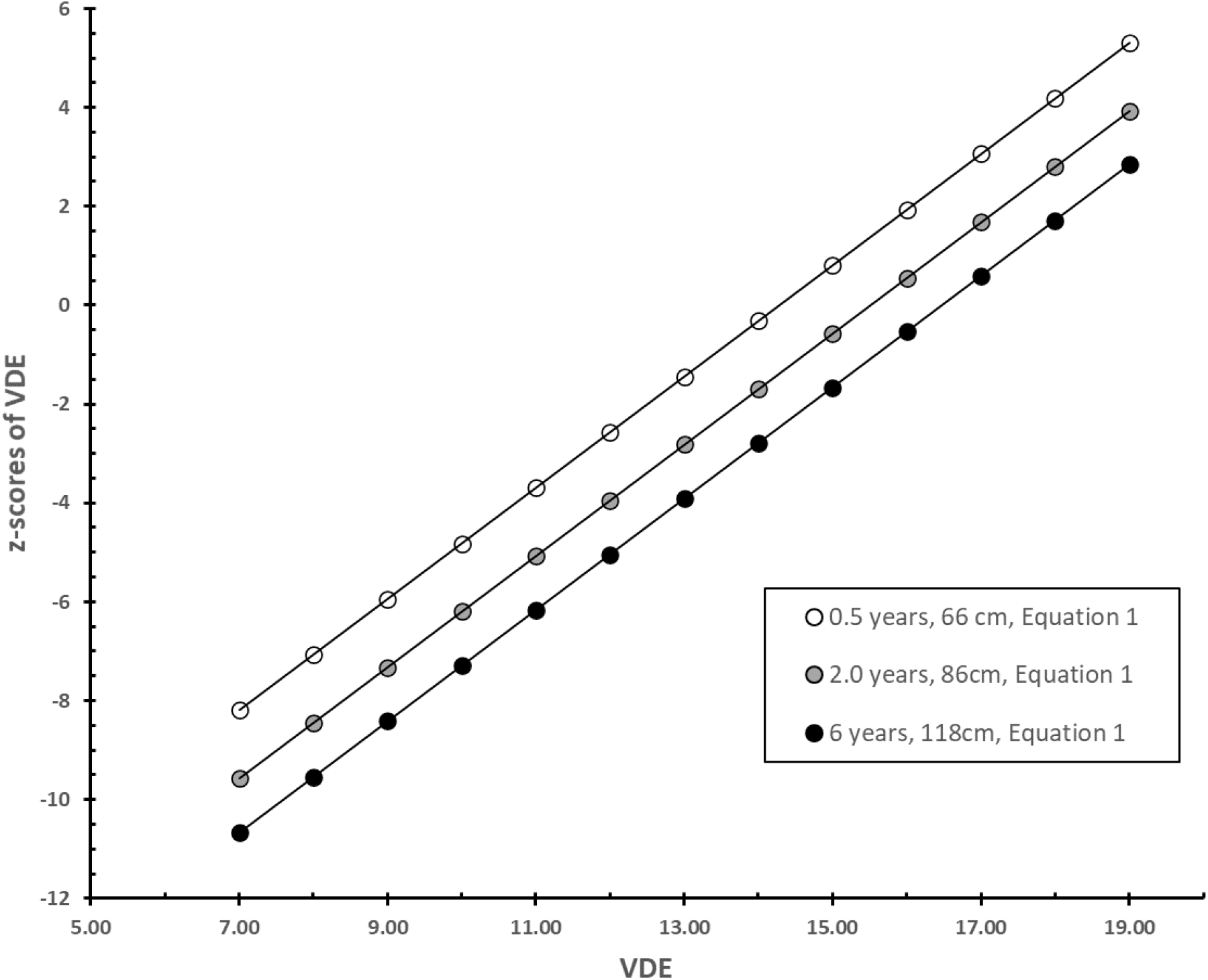
Ventilation distribution efficiency (VDE; FRC/CEV[%]) z-scores vs VDE with three different simulated values for heights and ages using Equation 1 of this paper

**Appendix Figure 4a.**
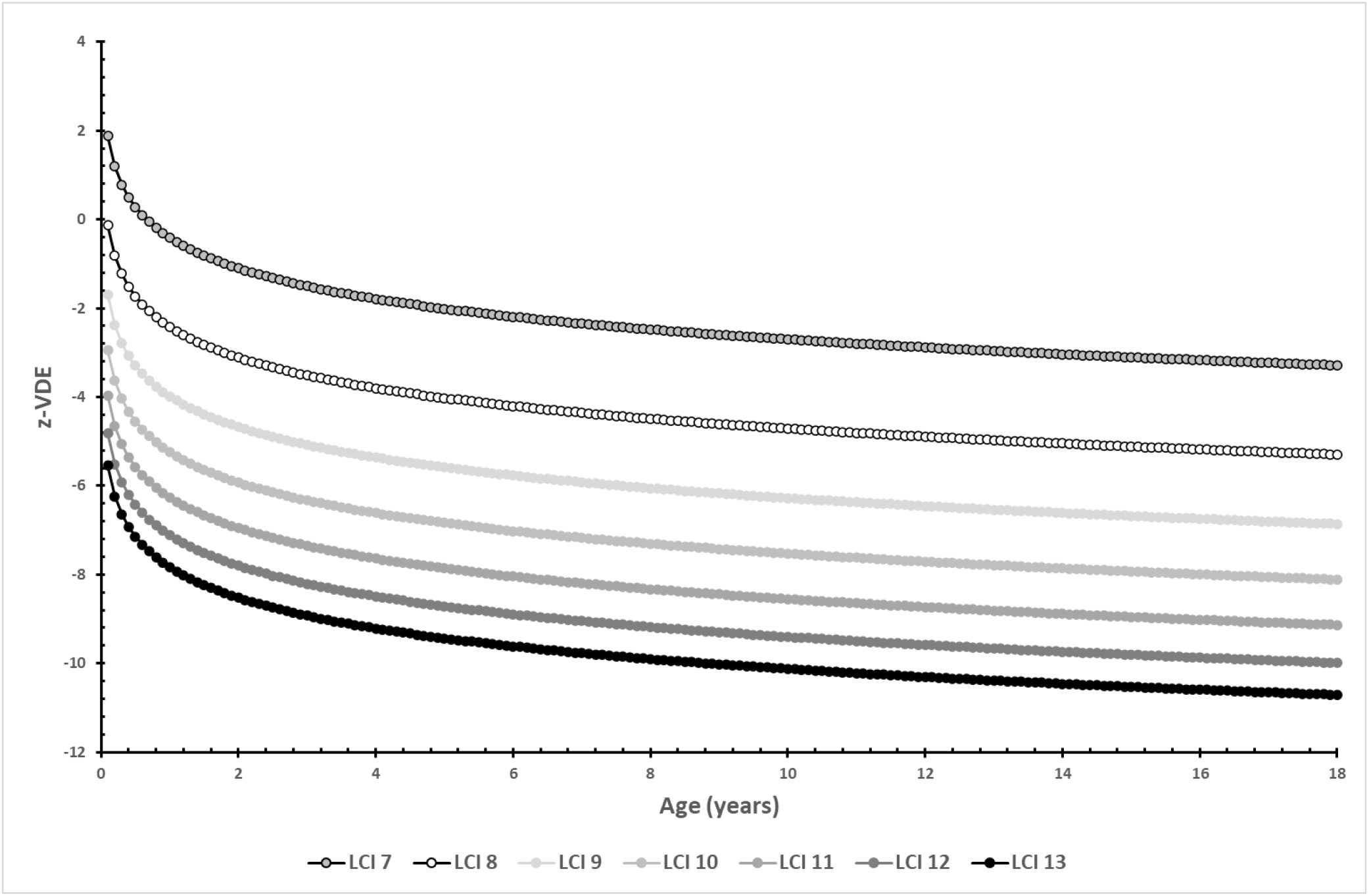
z-scores of ventilation distribution efficiency (VDE; FRC/CEV[%]) calculated across all ages from 0.1 to 18.0 years for fixed values of LCI 7, 8, 9, 10, 11, 12 and 13.

**Appendix Figure 4b.**
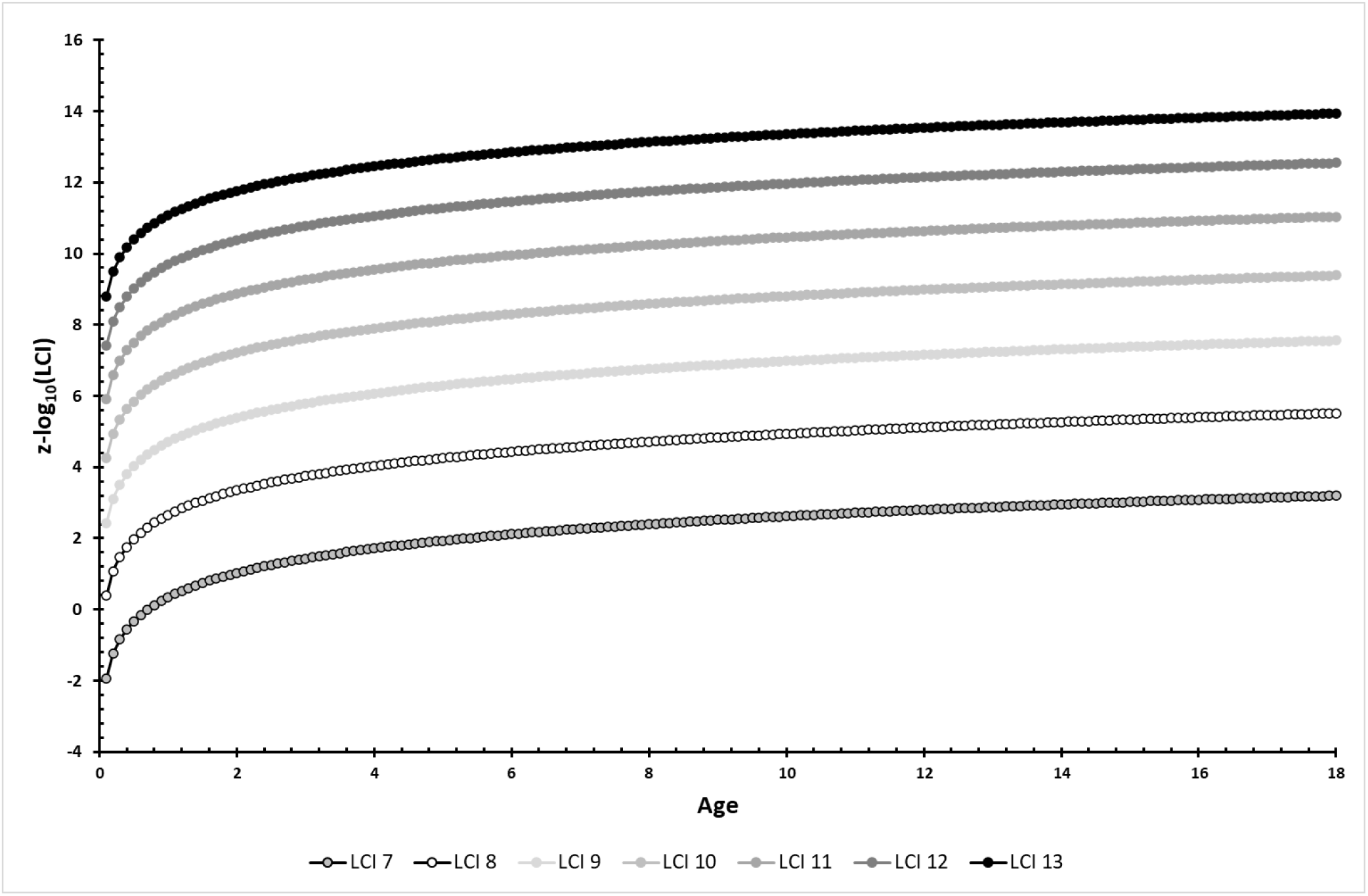
z-scores of log_10_(LCI) calculated across all ages from 0.1 to 18.0 years for fixed values of LCI 7, 8, 9, 10, 11, 12 and 13.

**Appendix Figure 4c.**
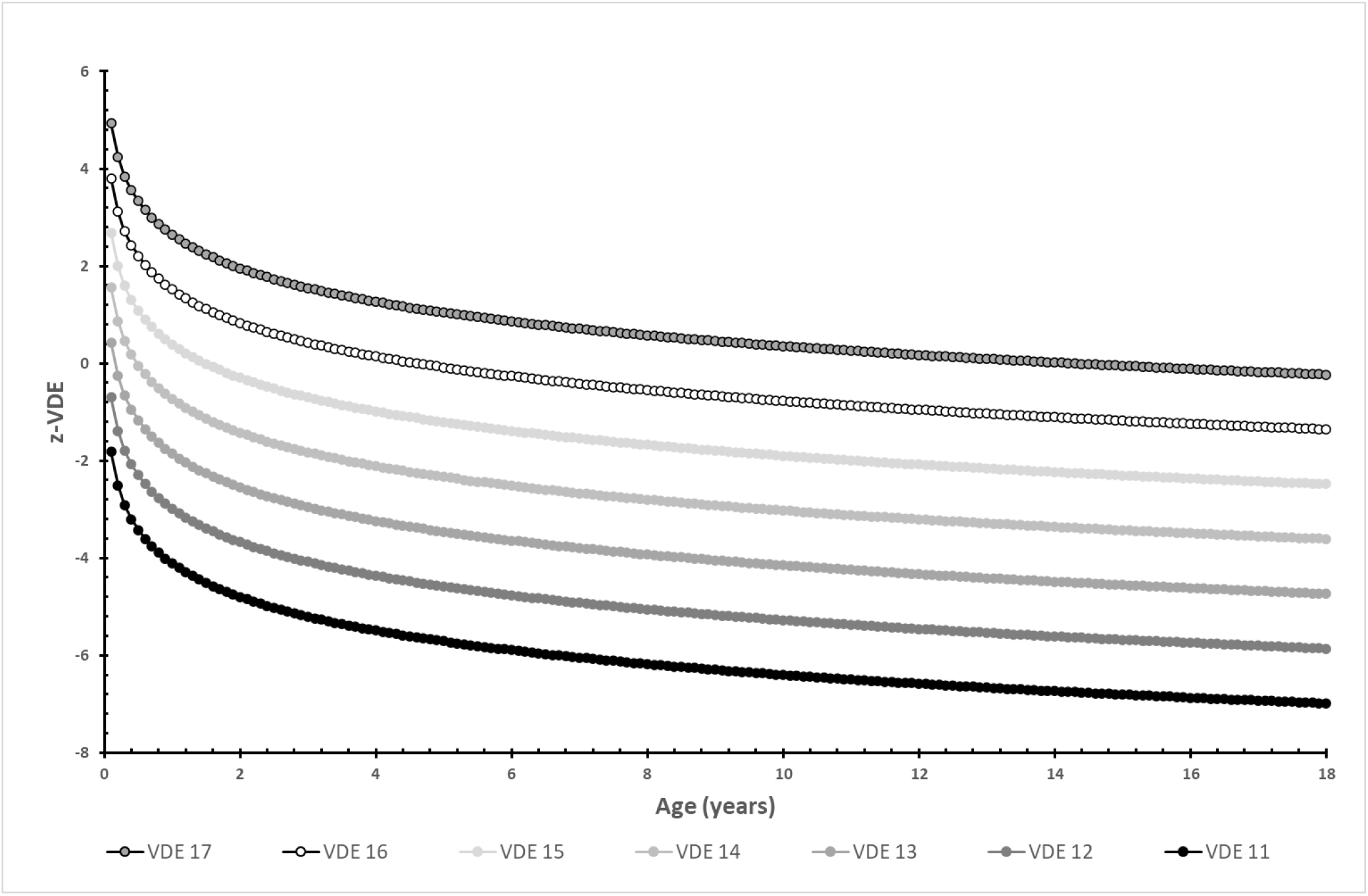
z-scores of ventilation distribution efficiency (VDE; FRC/CEV[%]) calculated across all ages from 0.1 to 18.0 years for fixed values of VDE 17, 16, 15, 14, 13, 12 and 11.

**Appendix Figure 4d.**
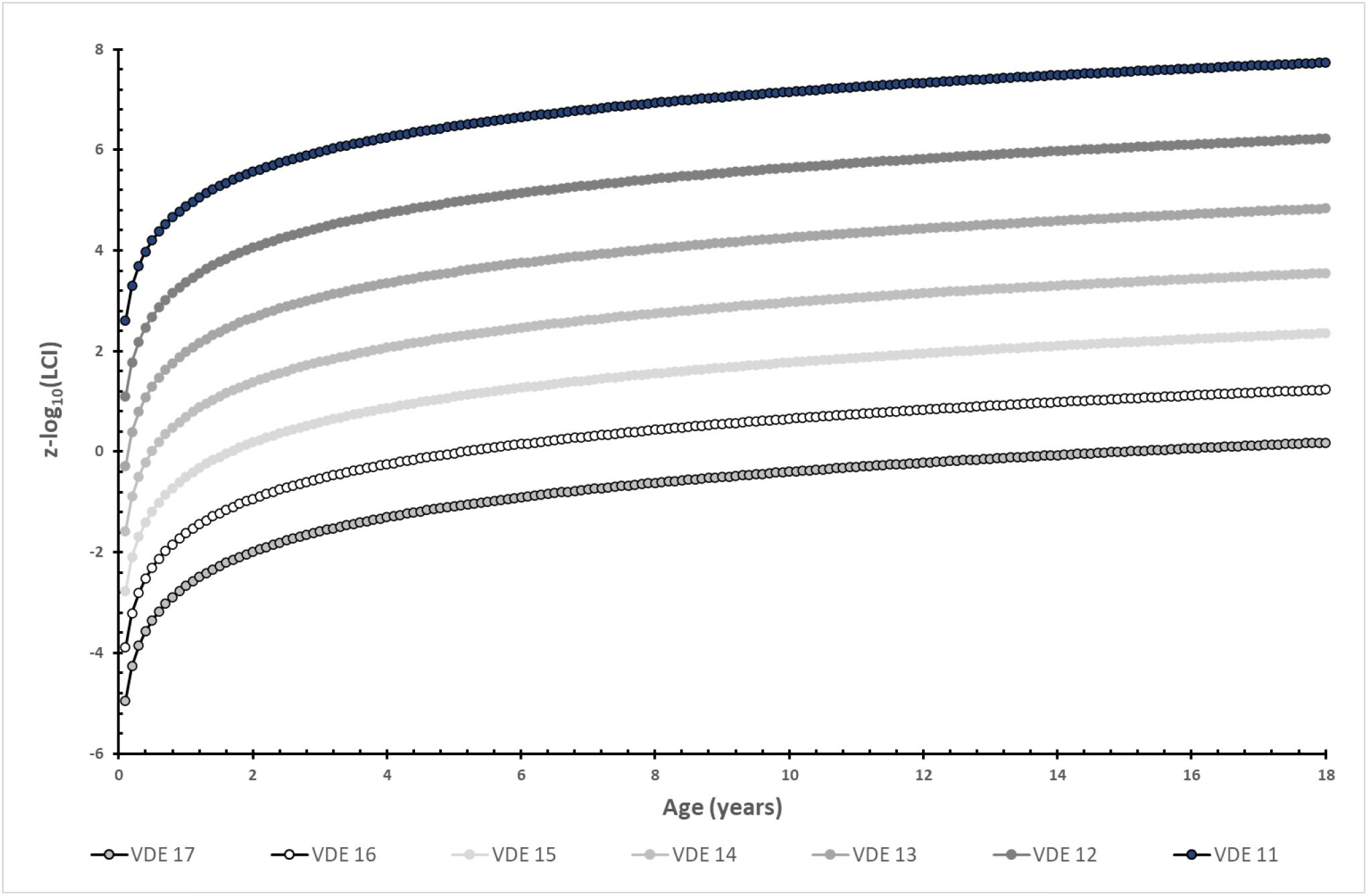
z-scores of log_10_(LCI) calculated across all ages from 0.1 to 18.0 years for fixed values of ventilation distribution efficiency (VDE; FRC/CEV[%]) 17, 16, 15, 14, 13, 12 and 11.

**Appendix Figure 5.**
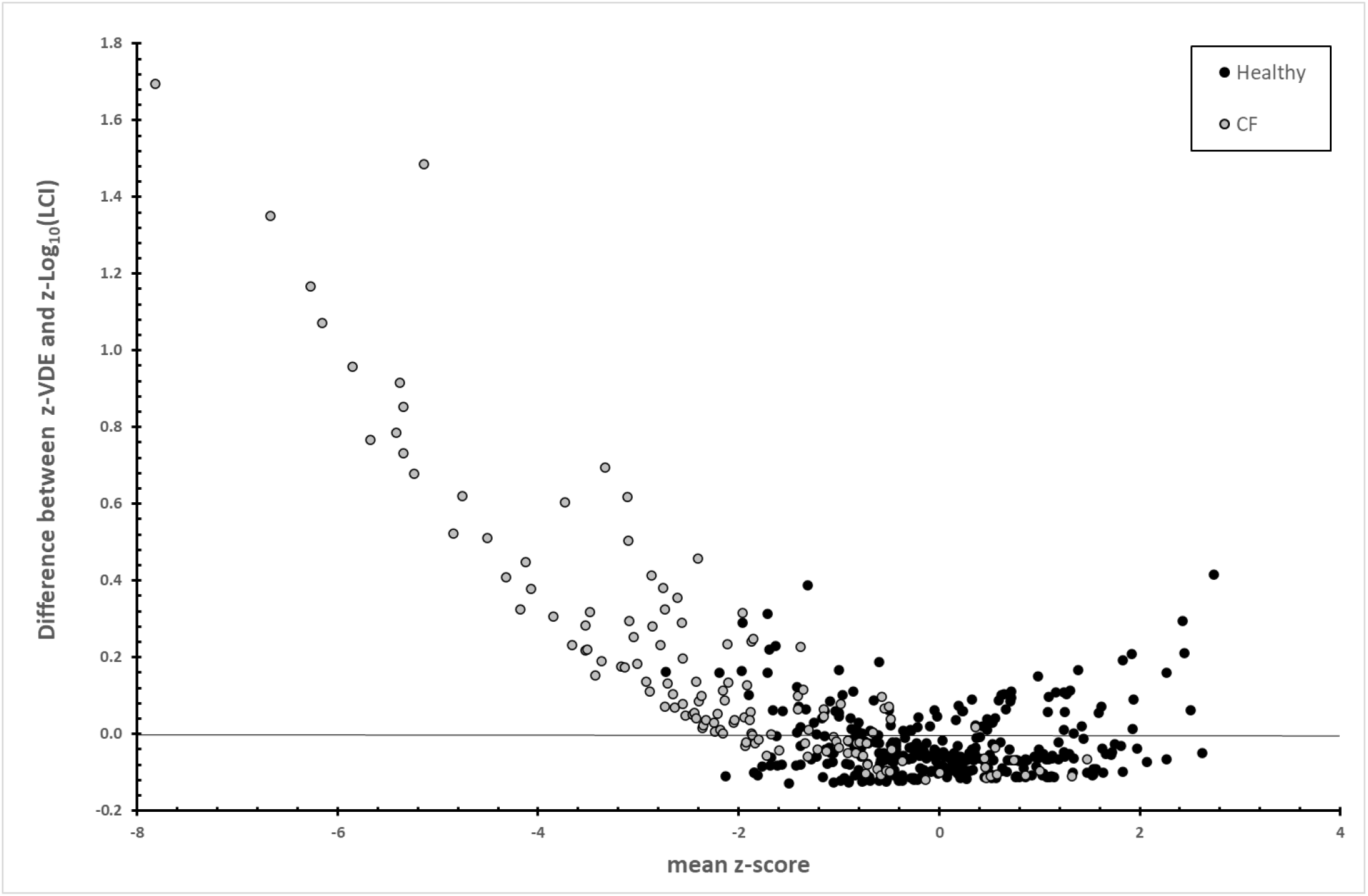
Differences in calculated z-scores of log_10_(LCI) and ventilation distribution efficiency (VDE; FRC/CEV[%]) for healthy children and children with CF, compared to the mean z-score.

## Notes

**Funding:** This study was supported by a Vertex Innovation Award; the Research Fund at Skaraborg Hospital, Skövde, Sweden; the SAKS Research Foundation, Uppsala, Sweden; the Children’s Lung Foundation (Denmark), and the research foundation at Copenhagen University Hospital, Rigshospitalet, Denmark.

### Competing Interest Statement

The authors have declared no competing interest.

### Funding Statement

This study was supported by a Vertex Innovation Award; the Research Fund at Skaraborg Hospital, Skovde, Sweden; the SAKS Research Foundation, Uppsala, Sweden; the Children's Lung Foundation (Denmark); and the research foundation at Copenhagen University Hospital, Rigshospitalet, Denmark.

## References

1. Robinson PD, Latzin P, Verbanck S, Hall GL, Horsley A, Gappa M, et al. Consensus statement for inert gas washout measurement using multiple- and single-breath tests. Eur Respir J. 2013 Mar 1;41(3):507–22.

2. Becklake MR. A New Index of the Intrapulmonary Mixture of Inspired Air. Thorax. 1952 Mar 1;7(1):111–6.

3. Ramsey KA, Rosenow T, Turkovic L, Skoric B, Banton G, Adams AM, et al. Lung Clearance Index and Structural Lung Disease on Computed Tomography in Early Cystic Fibrosis. Am J Respir Crit Care Med. 2015 Sep 11;193(1):60–7.

4. M. Sandvik R, Gustafsson PM, Lindblad A, Robinson PD, G. Nielsen K. Improved agreement between N2 and SF6 multiple-breath washout in healthy infants and toddlers with improved EXHALYZER D sensor performance. J Appl Physiol. 2021 Jul 1;131(1):107–18.

5. Schmidt MN, Sandvik RM, Voldby C, Buchvald FF, Jørgensen MN, Gustafsson P, et al. What it takes to implement regular longitudinal multiple breath washout tests in infants with cystic fibrosis. J Cyst Fibros. 2020 Nov 1;19(6):1027–8.

6. Sandvik RM, Gustafsson PM, Lindblad A, Buchvald F, Olesen HV, Olsen JH, et al. Contemporary N2 and SF6 multiple breath washout in infants and toddlers with cystic fibrosis. Pediatr Pulmonol. 2022;57(4):945–55.

7. Stahl M, Wielpütz MO, Graeber SY, Joachim C, Sommerburg O, Kauczor HU, et al. Comparison of Lung Clearance Index and Magnetic Resonance Imaging for Assessment of Lung Disease in Children With Cystic Fibrosis. Am J Respir Crit Care Med. 2016 Aug 30;rccm.201604-0893OC.

8. Stahl M, Wielpütz MO, Ricklefs I, Dopfer C, Barth S, Schlegtendal A, et al. Preventive Inhalation of Hypertonic Saline in Infants with Cystic Fibrosis (PRESIS). A Randomized, Double-Blind, Controlled Study. Am J Respir Crit Care Med. 2019 May 15;199(10):1238–48.

9. Thurlbeck WM. Postnatal growth and development of the lung. Am Rev Respir Dis. 1975 Jun;111(6):803–44.

10. Hislop AA, Wigglesworth JS, Desai R. Alveolar development in the human fetus and infant. Early Hum Dev. 1986 Feb;13(1):1–11.

11. Castile R, Filbrun D, Flucke R, Franklin W, McCoy K. Adult-type pulmonary function tests in infants without respiratory disease. Pediatr Pulmonol. 2000;30(3):215–27.

12. Stocks Janet. Infant respiratory function testing. New York: Wiley-Liss; 1996. 577 p. 13.

13. Lum S, Stocks J, Stanojevic S, Wade A, Robinson P, Gustafsson P, et al. Age and height dependence of lung clearance index and functional residual capacity. Eur Respir J. 2013 Jun 1;41(6):1371–7.

14. Lu Z, Dai R, Kowalik K, Dubeau A, Lefebvre DL, Balkovec S, et al. Newly developed multiple-breath washout reference equations from the CHILD Cohort Study: implications of poorly fitting equations. ERJ Open Res. 2021 Jan 25;7(1):00301–2020.

15. Gustafsson PM, Bengtsson L, Lindblad A, Robinson PD. The effect of inert gas choice on multiple breath washout in healthy infants: differences in lung function outcomes and breathing pattern. J Appl Physiol. 2017 Dec;123(6):1545–54.

16. Gustafsson PM, Robinson PD, Lindblad A, Oberli D. Novel methodology to perform sulfur hexafluoride (SF _6_)-based multiple-breath wash-in and washout in infants using current commercially available equipment. J Appl Physiol. 2016 Nov;121(5):1087–97.

17. Robinson PD, Latzin P, Ramsey KA, Stanojevic S, Aurora P, Davis SD, et al. Preschool Multiple-Breath Washout Testing. An Official American Thoracic Society Technical Statement. Am J Respir Crit Care Med. 2018 Mar 1;197(5):e1–19.

18. Arborelius M, Rosberg HE, Wiberg R. Multiple breath nitrogen dead space. Clin Physiol. 1988;8(6):561–76.

19. Cole TJ, Bellizzi MC, Flegal KM, Dietz WH. Establishing A Standard Definition For Child Overweight And Obesity Worldwide: International Survey. BMJ. 2000;320(7244):1240–3.

20. Tinggaard J, Aksglaede L, Sørensen K, Mouritsen A, Wohlfahrt-Veje C, Hagen CP, et al. The 2014 Danish references from birth to 20 years for height, weight and body mass index. Acta Paediatr. 2014;103(2):214–24.

21. Green K, Kongstad T, Skov M, Buchvald F, Rosthøj S, Marott JL, et al. Variability of monthly nitrogen multiple-breath washout during one year in children with cystic fibrosis. J Cyst Fibros. 2018 Mar 1;17(2):242–8.

22. Sandvik RM, Kongstad T, Green K, Voldby C, Buchvald F, Skov M, et al. Prospective longitudinal association between repeated multiple breath washout measurements and computed tomography scores in children with cystic fibrosis. J Cyst Fibros. 2021 Jul;20(4):632–40.

23. Svedberg M, Gustafsson PM, Robinson PD, Rosberg M, Lindblad A. Variability of lung clearance index in clinically stable cystic fibrosis lung disease in school age children. J Cyst Fibros. 2018 Mar;17(2):236–41.

24. Oude Engberink E, Ratjen F, Davis SD, Retsch-Bogart G, Amin R, Stanojevic S. Inter-test reproducibility of the lung clearance index measured by multiple breath washout. Eur Respir J. 2017;50(4).

25. Rigby RA, Stasinopoulos DM. Generalized additive models for location, scale and shape. J R Stat Soc Ser C Appl Stat. 2005;54(3):507–54.

26. Quanjer PH, Stanojevic S, Cole TJ, Baur X, Hall GL, Culver BH, et al. Multi-ethnic reference values for spirometry for the 3-95-yr age range: the global lung function 2012 equations. Eur Respir J. 2012 Dec;40(6):1324–43.

27. Robinson PD, Jensen R, Seeto RA, Stanojevic S, Saunders C, Short C, et al. Impact of cross-sensitivity error correction on representative nitrogen-based multiple breath washout data from clinical trials. J Cyst Fibros. 2022 May 1;21(3):e204–7.

28. Severinghaus JW, Stupfel M. Alveolar Dead Space as an Index of Distribution of Blood Flow in Pulmonary Capillaries. J Appl Physiol. 1957 May;10(3):335–48.

